# Genetic Risk Factors for ME/CFS Identified using Combinatorial Analysis

**DOI:** 10.1101/2022.09.09.22279773

**Authors:** Sayoni Das, Krystyna Taylor, James Kozubek, Jason Sardell, Steve Gardner

## Abstract

**Background:** Myalgic encephalomyelitis/chronic fatigue syndrome (ME/CFS) is a debilitating chronic disease that lacks known pathogenesis, distinctive diagnostic criteria, and effective treatment options. Understanding the genetic (and other) risk factors associated with the disease would begin to help to alleviate some of these issues for patients.

**Methods:** We applied both GWAS and the PrecisionLife combinatorial analytics platform to analyze ME/CFS cohorts from UK Biobank, including the Pain Questionnaire cohort, in a case-control design with 1,000 cycles of fully random permutation. Results from this study were supported by a series of replication and cohort comparison experiments, including use of disjoint Verbal Interview CFS, post-viral fatigue syndrome and fibromyalgia cohorts also derived from UK Biobank, and results compared for overlap and reproducibility.

**Results:** Combinatorial analysis revealed 199 SNPs mapping to 14 genes, that were significantly associated with 91% of the cases in the ME/CFS population. These SNPs were found to stratify by shared cases into 15 clusters (communities) made up of 84 high-order combinations of between 3-5 SNPs. *p*-values for these communities range from 2.3 × 10^−10^ to 1.6 × 10^−72^. Many of the genes identified are linked to the key cellular mechanisms hypothesized to underpin ME/CFS, including vulnerabilities to stress and/or infection, mitochondrial dysfunction, sleep disturbance and autoimmune development. We identified 3 of the critical SNPs replicated in the post-viral fatigue syndrome cohort and 2 SNPs replicated in the fibromyalgia cohort. We also noted similarities with genes associated with multiple sclerosis and long COVID, which share some symptoms and potentially a viral infection trigger with ME/CFS.

**Conclusions:** This study provides the first detailed genetic insights into the pathophysiological mechanisms underpinning ME/CFS and offers new approaches for better diagnosis and treatment of patients.

## Background

Myalgic encephalomyelitis/chronic fatigue syndrome (ME/CFS) is a debilitating chronic disease that presents with diverse symptoms including post-exertional malaise, chronic pain, and cognitive impairment^1^. It affects approximately 0.2% of the UK population^2^. There are currently no approved disease modifying therapies for ME/CFS, and patients are managed via prescription of drugs and other therapies for symptomatic relief, including pain relief, anti-depressants, and cognitive behavioural therapy^3^.

The breadth of symptoms and severities experienced by ME/CFS patients is likely indicative of the heterogeneous nature of the disorder, with a variety of metabolic, immunological, neuroendocrine and central nervous system dysfunctions underlying an individual patient’s pattern of onset and development of the disease. ME/CFS development has been associated with prior viral infection such as with Epstein-Barr Virus (EBV)^4^ and other pathogens^5,6,7,8^, however there is also evidence that stress and non-viral infection may also contribute to triggering ME/CFS onset^9^.

The multi-factorial spectrum of ME/CFS triggers and symptoms^10^ invites the question whether ME/CFS may represent multiple patient subgroups with a range of potentially overlapping underlying biological drivers. If so, better characterization of the etiology of disease in these subgroups may lead to improved understanding of ME/CFS and identification of personalized treatments that are most effective for specific subgroups.

Previous ME/CFS population studies have performed Genome-Wide Association Studies (GWAS) with the aim of identifying significant genetic factors underlying disease risk. While there is a demonstrable heritable component to the disease^11^, no significant single-gene association to ME/CFS has been identified using analysis of whole exome sequences, and given the limited statistical power associated with the small ME/CFS genetic datasets available, GWAS approaches have been unable to detect disease-associated SNPs that exhibit sufficiently large effect sizes across the whole of the patient population^12^.

ME/CFS is clearly not a simple monogenic disease caused by single nucleotide variants (SNVs) with large effect sizes but by the complex interactions of many genetic, epidemiological and environmental factors that GWAS-based approaches are not able to fully identify. This requires a different analytical approach.

### Combinatorial Analysis

Although GWAS has helped to transform the treatment of many relatively monogenic diseases by revealing clinically relevant single SNP genetic associations, it has been less successful in complex, chronic diseases. These are more polygenic and heterogeneous with patients presenting in a spectrum, and they may include high-resolution signals such as disease-associated variants occurring within linkage disequilibrium (LD) blocks^13,14^.

Notably, inclusion of patients with different etiologies under the same “case” classification weakens SNP-disease associations in GWAS, causing the method to potentially overlook the genetic variants responsible for disease in subsets of the population. More fundamentally, GWAS is not designed to detect epistatic and other non-linear effects caused by the interactions of multiple variants. As such it struggles to identify variants that are strongly associated with different patient subgroups in a heterogeneous patient population with multiple diverse disease etiologies that may be further influenced by non-linear interactions across and between multiple genes and transcription/expression control regions.

This however is exactly the challenge presented by ME/CFS and other complex, chronic diseases. Understanding of how the range of disease etiologies affects different patient subgroups requires the identification of combinations of SNPs (and other clinical, transcriptomic and/or epidemiological or environmental features) that together are co-associated with a specific phenotype.

The PrecisionLife combinatorial analysis platform enables hypothesis-free identification of such high-order combinatorial multi-modal features (known as disease signatures) at scale on modest computational hardware. These combinatorial disease signatures capture both linear and non-linear effects of genetic and molecular interaction networks in a way that is complementary to GWAS analysis^15^.

The combinatorial approach is more sensitive than GWAS, enabling identification of novel genetic associations and mechanisms that may only be relevant to a subgroup of patients, leading to more validated associations than GWAS when analyzing the same datasets. This approach has been validated in multiple disease studies both by the authors and collaborators, in some cases using *in vitro* and *in vivo* disease assays to demonstrate novel target genes’ disease modification potential, and in others by the presence in pharmaceutical companies’ R&D pipelines of drug programs targeting mechanisms that were identified by combinatorial analysis, but which could not be found using GWAS on available patient datasets^16,17,18^.

For example, using combinatorial analysis we were first to report the association of 156 loci and 68 genes with the risk of developing severe COVID-19^19^. This analysis was run on just 725 patients and 1,450 controls from UK Biobank, and contrasts with the 11 and 13 loci discovered using a GWAS approach respectively by 23andMe (16,500 patients/controls)^20^ and COVID-19 HGI consortium (over 2,000,000 patient/controls)^21^ in similar studies. Of the 68 genes that we reported, 48 were subsequently associated with the disease by other groups using methods including single-cell analysis and transcriptomic profiling (unpublished literature analysis - June 2022).

## Materials and Methods

We analyzed genotype data from 2,382 patients reporting an ME/CFS diagnosis in the UK Biobank Pain Questionnaire^22^ matched against 4,764 controls in a case:control study design in the PrecisionLife platform.

### Data Sources

ME/CFS patients with a (self-reported) clinical diagnosis in UK Biobank’s Pain Questionnaire (Data-field 120010) were identified, of whom over 90% were of European genetic ancestry (Figure 10 in Supplementary Data). Given this proportion, only ME/CFS patients of European genetic ancestry were selected as the case cohort for this study. To ensure properly characterized control subjects, individuals were selected who had no evidence in the Hospital Episodes Statistics (HES), primary care, or self-reported data fields indicating diagnoses of chronic fatigue, post-exertional malaise, post-viral fatigue syndrome or myalgia (see Table 9 in Supplementary Data). To avoid potential confounding, controls meeting these criteria were matched by genetic ancestry and gender against the cases in a 2:1 ratio (and this was repeated with a separate 4:1 ratio study). Because of the narrow age range of UK Biobank participants, strict age matching was not necessary, as cases and controls exhibit very similar age distributions (Figure 12a in Supplemental Data).

**Table 1:** Summary of thresholds used in the mining phase of the PrecisionLife combinatorial analysis on the Pain Questionnaire cohort.

| Layer | Minimum Z-Score | Minimum Cases (%) | Minimum Risk Ratio | No. of Reported Disease Signatures |
| --- | --- | --- | --- | --- |
| 1 (singleton) | 1.96 | 5% | 2.00 | 0 |
| 2 (pair) | 2.85 | 5% | 2.00 | 184 |
| 3 (triple) | 3.83 | 5% | 2.34 | 45,711 |
| 4 (quadruple) | 4.21 | 5% | 3.10 | 49,894 |
| 5 (quintuple) | 4.33 | 5% | 3.97 | 44,426 |

**Table 2:** Summary of the results of PrecisionLife combinatorial analysis run on the Pain Questionnaire cohort.

|  |  |
| --- | --- |
| **Validated Disease Signatures (SNP Combinations)** | 84 |
| **Total Disease Associated SNPs in Disease Architecture** | 199 |
| **Critical SNPs** | 25 |
| **Prioritized Genes** | 14 |
| **Cases represented by Disease Architecture (%)** | 91% |

**Table 3:** Example of one of the combinatorial disease signatures contributing to Community 1 identified by the PrecisionLife combinatorial analysis of the Pain Questionnaire cohort. **Bold** text indicates the critical (RF-scored) SNPs (and the genes to which they are mapped) in this signature.

| SNP ID/Genotype | Mapped Genes | # Cases (t=2,382) | # Controls (t=4,764) |
| --- | --- | --- | --- |
| rs58634322 / 0 | **AC009486.1** | 120 | 48 |
| rs7731560 / 2 |  |  |  |
| rs77821266 / 0 | AP001978.1 |  |  |
| **rs237475 / 1** | **KCNB1/** |  |  |
|  | **AL035685.1** |  |  |
| rs2904106 / 0 | **ATP9A** |  |  |

**Table 4:** Disease signatures, SNPs and case count associated with communities identified in the Pain Questionnaire cohort.

| Community Number | Disease Signatures | SNPs | Case Count (t=2,382) | Control Count (t=4,764) | Chi Square p-value | Odds Ratio |
| --- | --- | --- | --- | --- | --- | --- |
| 1 | 6 | 8 | 144 | 102 | $2.56 \times 10^{-17}$ | 2.94 |
| 2 | 1 | 5 | 153 | 72 | $8.30 \times 10^{-29}$ | 4.47 |
| 3 | 2 | 5 | 257 | 203 | $5.13 \times 10^{-26}$ | 2.72 |
| 4 | 4 | 9 | 419 | 371 | $2.10 \times 10^{-35}$ | 2.53 |
| 5 | 2 | 4 | 228 | 197 | $8.46 \times 10^{-20}$ | 2.45 |
| 6 | 12 | 20 | 744 | 719 | $5.36 \times 10^{-57}$ | 2.56 |
| 7 | 7 | 16 | 408 | 315 | $1.18 \times 10^{-43}$ | 2.92 |
| 8 | 4 | 11 | 343 | 231 | $2.86 \times 10^{-44}$ | 3.30 |
| 9 | 17 | 38 | 648 | 503 | $1.60 \times 10^{-72}$ | 3.17 |
| 10 | 6 | 17 | 425 | 376 | $5.24 \times 10^{-36}$ | 2.53 |
| 11 | 6 | 21 | 469 | 348 | $5.55 \times 10^{-54}$ | 3.11 |
| 12 | 3 | 10 | 142 | 136 | $2.34 \times 10^{-10}$ | 2.16 |
| 13 | 6 | 15 | 282 | 207 | $5.08 \times 10^{-32}$ | 2.96 |
| 14 | 4 | 14 | 348 | 193 | $1.23 \times 10^{-56}$ | 4.05 |
| 15 | 2 | 6 | 170 | 89 | $5.94 \times 10^{-29}$ | 4.04 |

**Table 5:** Genes and communities identified in the Pain Questionnaire cohort associated with phenotypic and clinical features.

| Gene | SNP (Most severe variant consequence – VEP) | SNP Minor Allele Frequency | Gene association with Phenotypic or Clinical feature (p-value before multiple testing correction) | Community | Community association with Phenotypic or Clinical feature (p-value before multiple testing correction) | Gene association replicated in disjoint Verbal Interview Cohort |
| --- | --- | --- | --- | --- | --- | --- |
| *S100PBP* | rs41306603 (3 prime UTR variant) | 0.052 | Males ($p=0.022$) | 1 | Males ($p=0.016$) | No |
| *ATP9A* | rs2904106 (intron variant) | 0.271 | Males ($p=0.016$) | 1 | Males ($p=0.016$) | Yes |
| *KCNB1* | rs237475 (intron variant) | 0.472 | Males ($p=0.016$) | 1 | Males ($p=0.016$) | No |
| *CLOCK* | rs6832769 (3 prime UTR variant) | 0.362 | Fibromyalgia, ICD-10: M79.7 ($p=0.026$);<br>Other soft tissue disorders, ICD-10: M79 ($p=0.044$) | 2 | Fibromyalgia, ICD-10: M79.7 ($p=0.026$);<br>Other soft tissue disorders, ICD-10: M79 ($p=0.044$) | No |
| *SLC15A4* | rs2398428 (intergenic variant) | 0.269 |  | 3 |  | No |
| *TMEM232* | rs58264436 (intergenic variant) | 0.394 |  | 6 |  | No |
| *GPC5* | rs16947237 (intron variant) | 0.155 |  | 7 |  | No |
| *PHACTR2* | rs9403525 (intron variant) | 0.448 | Thyroiditis, ICD10-10: E06 ($p=0.033$) | 8 | Thyroiditis, ICD10-10: E06 ($p=0.033$) | No |
| *AKAP1* | rs3785477 (intron variant) | 0.398 |  | 9 |  | No |
| *USP6NL* | rs2499908 (intergenic variant) | 0.235 |  | 12 |  | No |
| *CDON* | rs73021223 (intron variant) | 0.093 | Illness, injury, bereavement, stress in last 2 years 2014+ ($p=0.023$) | 13 | | No |
| *INSR* | rs59165976 (intron variant) | 0.388 |  | 14 |  | No |
| *SLC6A11* | rs2304725 (synonymous variant) | 0.319 | Phenylalanine levels in plasma ($p=0.022$);<br>Illness, injury, bereavement, stress in | 15 | Illness, injury, bereavement, stress in last 2 years 2019+ ($p=0.003$) | Yes |

| | | | last 2 years 2019+<br>($p=0.003$) | | | |
| --- | --- | --- | --- | --- | --- | --- |
| *SULF2* | rs56218501/<br>Affx-16805420<br>(missense variant) | 0.213 | Phenylalanine levels in<br>plasma ($p=0.022$);<br>Illness, injury,<br>bereavement, stress in<br>last 2 years 2019+<br>($p=0.003$) | 15 | Illness, injury, bereavement,<br>stress in last 2 years 2019+<br>($p=0.003$) | No |

**Table 6:** ME/CFS Pain Questionnaire critical SNPs and genes that were replicated in three disjoint fatigue-associated cohorts – CFS Verbal Interview, Post-viral syndrome and Fibromyalgia.

| Critical SNP (Gene) in ME/CFS Pain Questionnaire cohort | CFS Verbal Interview cohort | Post-viral syndrome cohort | Fibromyalgia cohort |
| --- | --- | --- | --- |
| rs2304725 (*SLC6A11*) | ☑ |  |  |
| rs2904106 (*ATP9A*) | ☑ |  |  |
| rs9444564 | ☑ |  |  |
| rs10420798 | ☑ |  | ☑ |
| rs11695478 | ☑ |  |  |
| Affx-16805420 (*SULF2*) |  | ☑ |  |
| rs12530627 |  | ☑ |  |
| rs2499908 (*USP6NL*) |  | ☑ | ☑ |

**Table 7:** Genes and communities identified in the Pain Questionnaire cohort and their proposed mechanism of action (MoA) in ME/CFS development (accounting for eQTL directionality).

| Gene | Community | MoA Category | MoA Evidence |
| --- | --- | --- | --- |
| *S100PBP* | 1 | Viral/Bacterial Susceptibility | Identified as a differentially expressed gene (DEG) in response to RSV infection<sup>31</sup> |
| *ATP9A* | 1 | Metabolic | Exhibits P4-ATPase activity in beta cells and *ATP9A* knockdown results in reduction in glucose-stimulated insulin release<sup>32</sup> |
|  |  | Autoimmune | Splicing region in *ATP9A* associated with multiple sclerosis risk in whole-exome sequencing analysis<sup>33</sup> |
| *KCNB1* | 1 | Metabolic | Regulates insulin exocytosis from pancreatic beta cells<sup>34</sup> |
| *CLOCK* | 2 | Sleep | Key regulator of circadian rhythm<sup>35</sup> |
|  |  | Metabolic | Disruption to circadian clock results in mitochondrial dysfunction and insulin resistance<sup>36,37</sup> |
| *SLC15A4* | 3 | Autoimmune | *SLC15A4* mediates type 1 interferon production and generation of autoantibodies in systemic lupus erythematosus (SLE)<sup>38</sup> |
|  |  | Metabolic | Knock down of *SLC15A4* decreases mitochondrial membrane potential and impairs AMPK activation<sup>39,40</sup> |
| *TMEM232* | 6 | Autoimmune | The promoter region of *TMEM232* is differentially methylated in a study of multiple sclerosis patients<sup>41</sup> |
| *GPC5* | 7 | Autoimmune | Polymorphisms in *GPC5* have been associated with multiple sclerosis risk in GWAS<sup>42,43</sup> |
| *PHACTR2* | 8 | Autoimmune | Identified as a multiple sclerosis risk gene in a GWA analysis<sup>44</sup> |
| *AKAP1* | 9 | Metabolic | *AKAP1* is an AMPK substrate that regulates mitochondrial respiration<sup>45</sup> |
|  |  | Viral/Bacterial Susceptibility | *AKAP1*<sup>-/-</sup> mice exhibit increased inflammation and immune cell infiltration in lungs<sup>46</sup> |
| *USP6NL* | 12 | Viral/Bacterial Susceptibility | Plays a role in HSV-1 virion assembly<sup>47</sup> |
| *CDON* | 13 | Metabolic | Cdon depletion results in impaired muscle regeneration and increased cell stress<sup>48</sup> |
|  |  | Autoimmune | Cdon is involved in differentiation and myelination of oligodendrocytes<sup>49</sup> |
|  |  | Viral/Bacterial Susceptibility | GWAS indicates *CDON* plays a role in complicated *Staphylococcus aureus* bacteremia<sup>50</sup> |
| *INSR* | 14 | Metabolic | Receptor tyrosine kinase which mediates insulin signalling<sup>51</sup> |
| *SULF2* | 15 | Viral/Bacterial Susceptibility | Genome-wide environmental interaction study indicates an association between a locus in *SULF2* and HSV-1 seropositivity and depression<sup>52</sup> |
|  |  | Metabolic | Overexpression of *SULF2* observed in subjects with type 2 diabetes and obesity<sup>53</sup> |
| *SLC6A11* | 15 | Sleep | Associated with depression, sleep disturbance and neuropathic pain in mouse models<sup>54,55</sup> |
|  |  | Autoimmune | Expression decreased in the hippocampus of mouse model of multiple sclerosis (EAE)<sup>56</sup> |

**Table 8:** Genes identified in the Pain Questionnaire cohort with their tractability as drug targets using annotations from OpenTargets^110^.

| Gene | Small Molecule Tractable | Small Molecule Clinical | Antibody Tractable | Existing Drugs | Development Compounds | PDB Structures |
| --- | --- | --- | --- | --- | --- | --- |
| *S100PBP* | 0 | 0 | 0 | 0 | 0 | 0 |
| *ATP9A* | 0 | 0 | 1 | 0 | 0 | 0 |
| *KCNB1* | 0.3 | 1 | 1 | 7 | 28 | 0 |
| *CLOCK* | 0 | 0 | 0 | 0 | 0 | 2 |
| *SLC15A4* | 0 | 0 | 1 | 0 | 0 | 0 |
| *TMEM232* | 0 | 0 | 0 | 0 | 0 | 0 |
| *GPC5* | 0 | 0 | 0.3 | 0 | 0 | 0 |
| *PHACTR2* | 0 | 0 | 0.3 | 0 | 0 | 0 |
| *AKAP1* | 0 | 0 | 0 | 0 | 0 | 0 |
| *USP6NL* | 0 | 0 | 0.3 | 0 | 0 | 0 |
| *CDON* | 0 | 0 | 0.3 | 0 | 0 | 3 |
| *INSR* | 1 | 0.7 | 0.3 | 20 | 891 | 62 |
| *SULF2* | 0 | 0 | 0 | 0 | 0 | 0 |
| *SLC6A11* | 0.3 | 0 | 0.3 | 6 | 126 | 0 |

**Table 9:** Control selection criteria for the ME/CFS Pain Questionnaire cohort

| Data Type | ICD-10 / Data Field ID | Description |
| --- | --- | --- |
| HES | G93.3 | Postviral fatigue syndrome |
| HES | R53.x | Malaise and fatigue |
| HES | G47.x | Sleep disorders |
| HES | M79.7 | Fibromyalgia |
| Self-reported | 120009 | Ever had fibromyalgia syndrome? |
| Self-reported | 120010 | Ever had Chronic Fatigue Syndrome or Myalgic Encephalitis? |
| Self-reported | 120120 | Exercise brings on fatigue |
| Self-reported | 120122 | Fatigue interferes with physical functioning |
| Self-reported | 120123 | Fatigue causes frequent problems |
| Self-reported | 120124 | Fatigue prevents sustained physical functioning |
| Self-reported | 120125 | Fatigue interferes with carrying out certain duties or responsibilities |
| Self-reported | 120126 | Fatigue is among three most disabling symptoms |
| Self-reported | 120127 | Fatigue interferes with work, family or social life |

**Figure 1.**
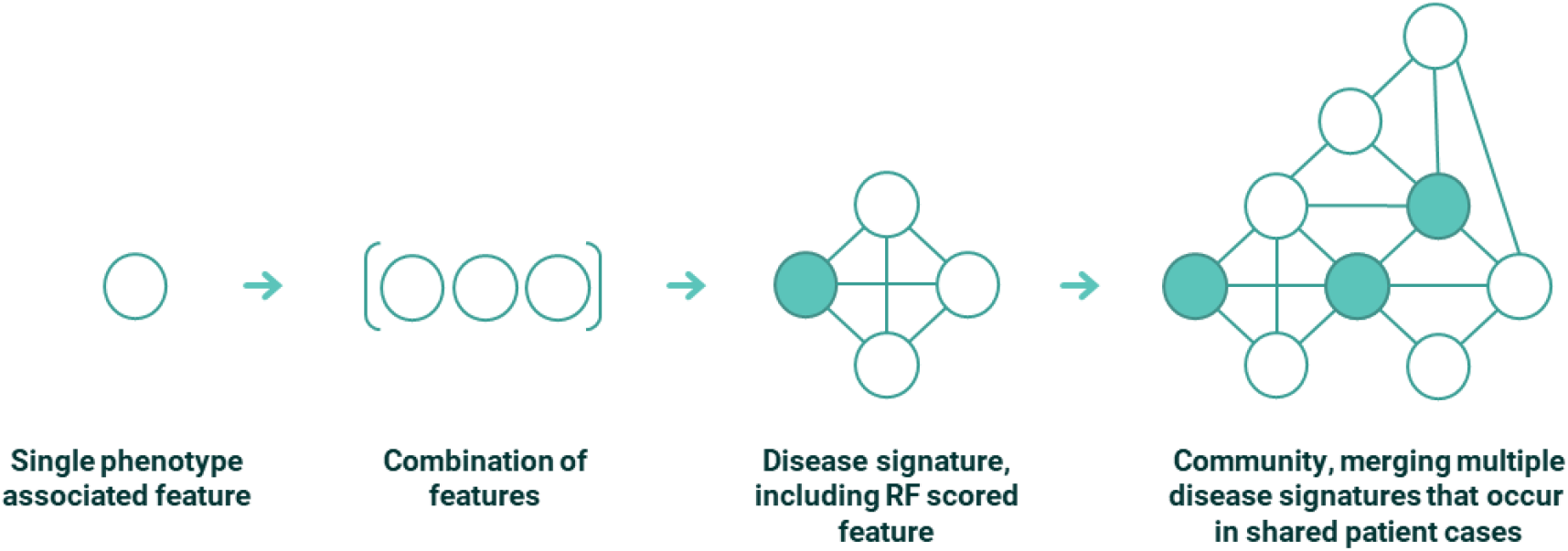
Conceptual representation of features, combinations, disease signatures and communities used to build up the disease architecture in the PrecisionLife combinatorial methodology. In the case of the ME/CFS study all features were SNP genotypes, but other feature types, e.g., a patient’s expression level of a specific protein, medication history or their eosinophil level, can also be used.

**Figure 2.**
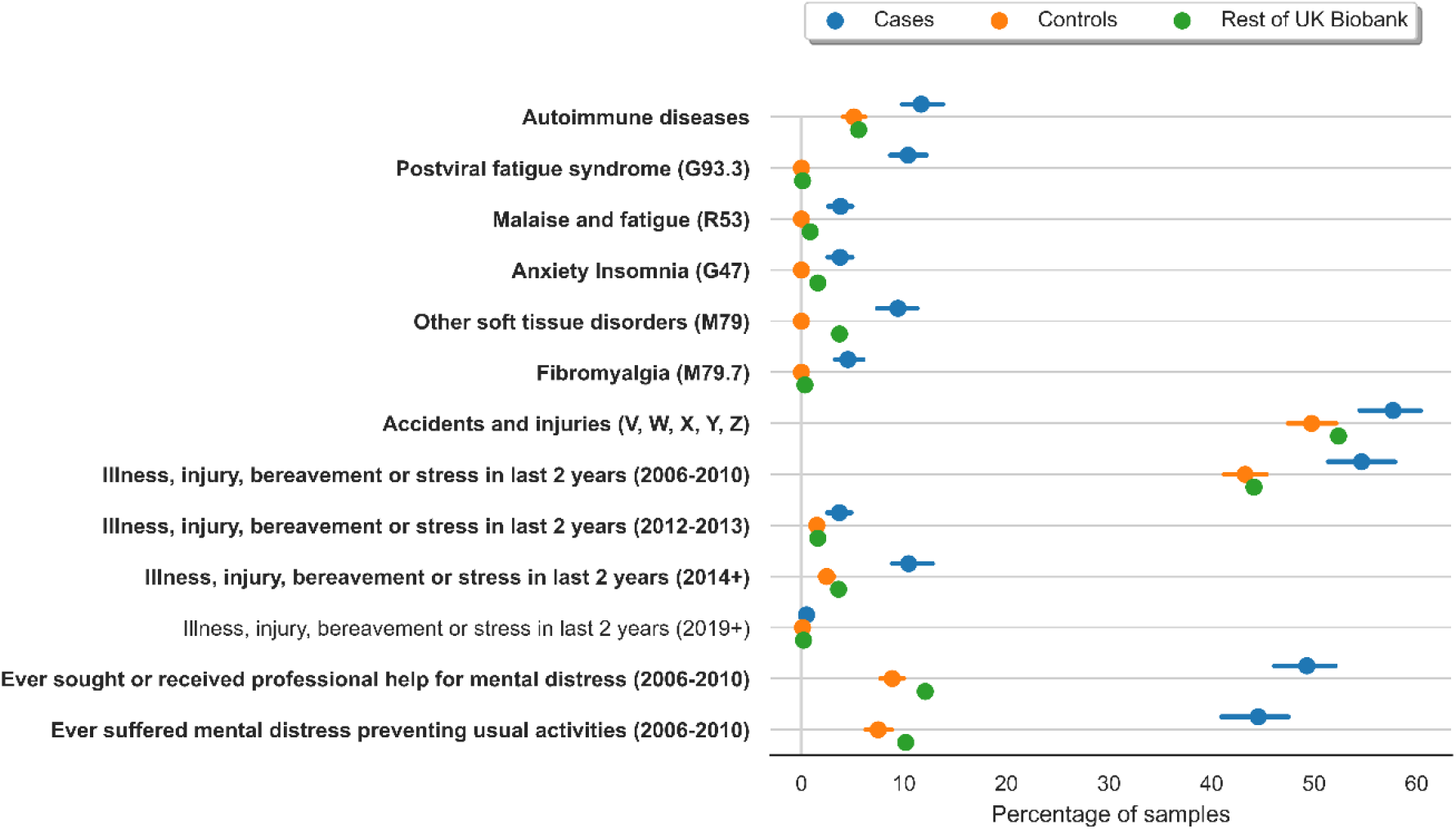
Forest plot showing percentage of individuals in cases, controls and rest of the individuals in UK Biobank who report each covariate along with 95% confidence interval generated using bootstrapping for 1,000 iterations. **Bold** covariate label indicates *p*<0.001, regular label indicates *p*<0.01.

**Figure 3:**
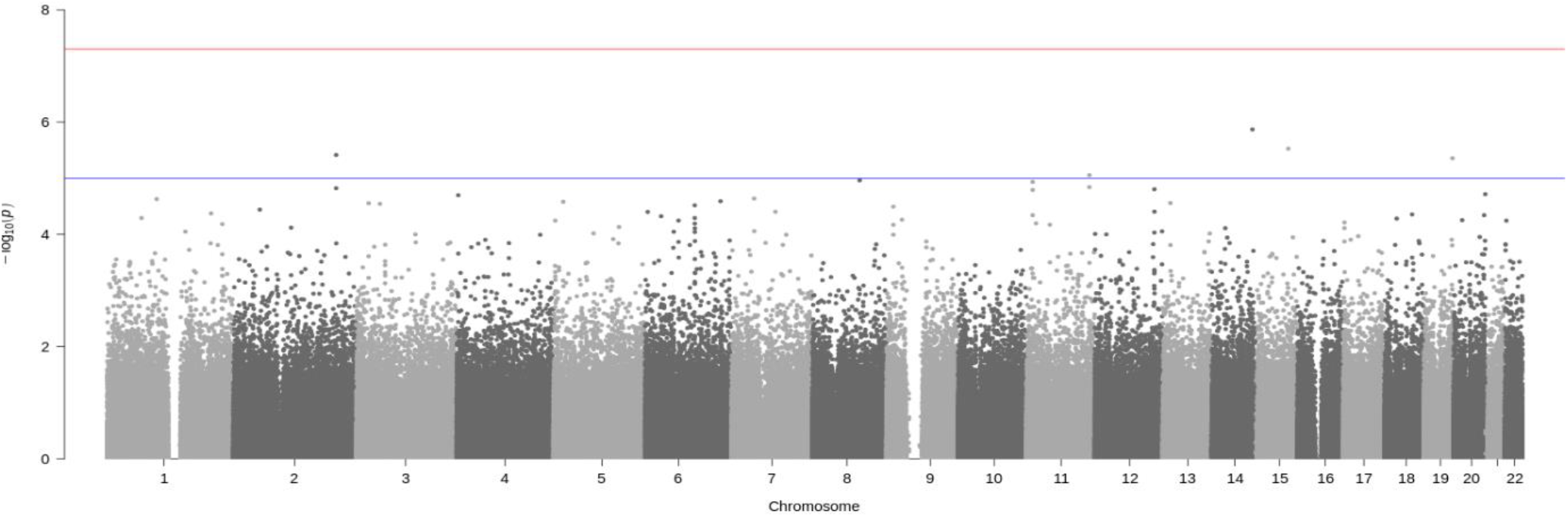
Manhattan plot generated using PLINK of genome-wide *p*-values of association for the Pain Questionnaire cohort (n = 7,146 where cases=2,382 and controls=4,764). The horizontal blue and red lines represent the genome-wide significance values of *p*<1 × 10^−5^ and *p*<5 × 10^−8^ respectively.

**Figure 4:**
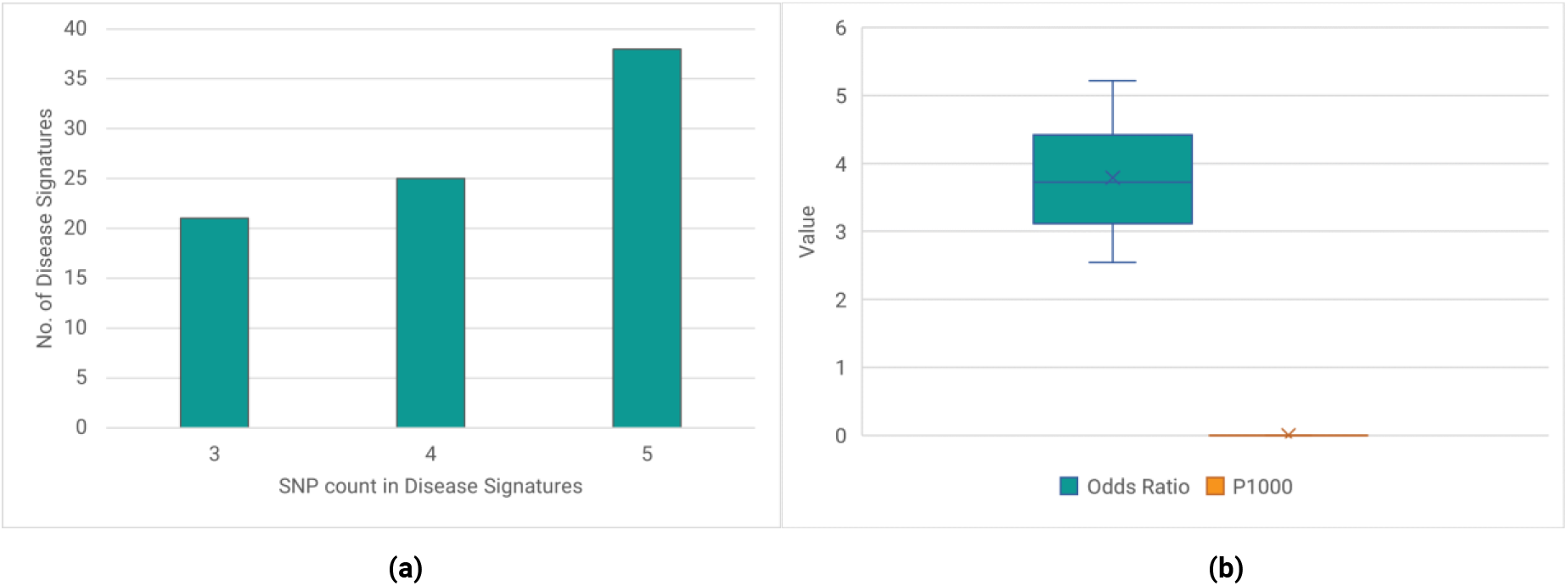
(a) Distribution of the combinatorial order of the 84 validated combinatorial disease signatures identified in the Pain Questionnaire cohort – i.e., 3 = signatures containing 3 co-associated SNPs. (b) Boxplot showing distribution of odds ratio and P1000 associated with 84 disease signatures identified in the Pain Questionnaire cohort.

**Figure 5:**
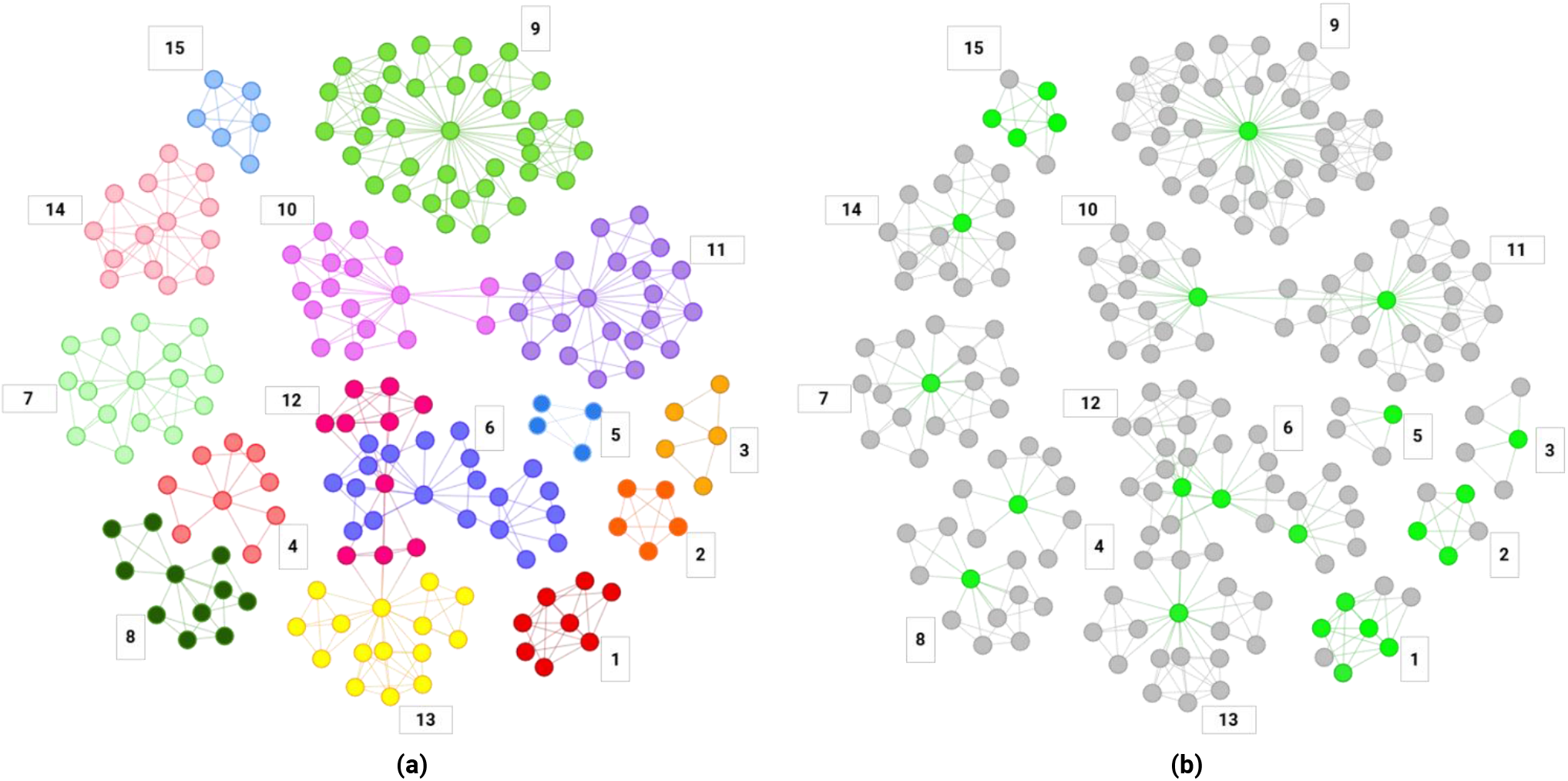
(a) Disease architecture diagram demonstrating the 15 communities of SNPs that make up the structure of the Pain Questionnaire patient sub-populations generated by the PrecisionLife platform. Each circle represents a disease-associated SNP genotype, edges represent their co-association in patients in disease signature(s), and colours represent distinct patient sub-populations. the same disease architecture view coloured to show the critical SNPs associated with each community (light green)

**Figure 6:**
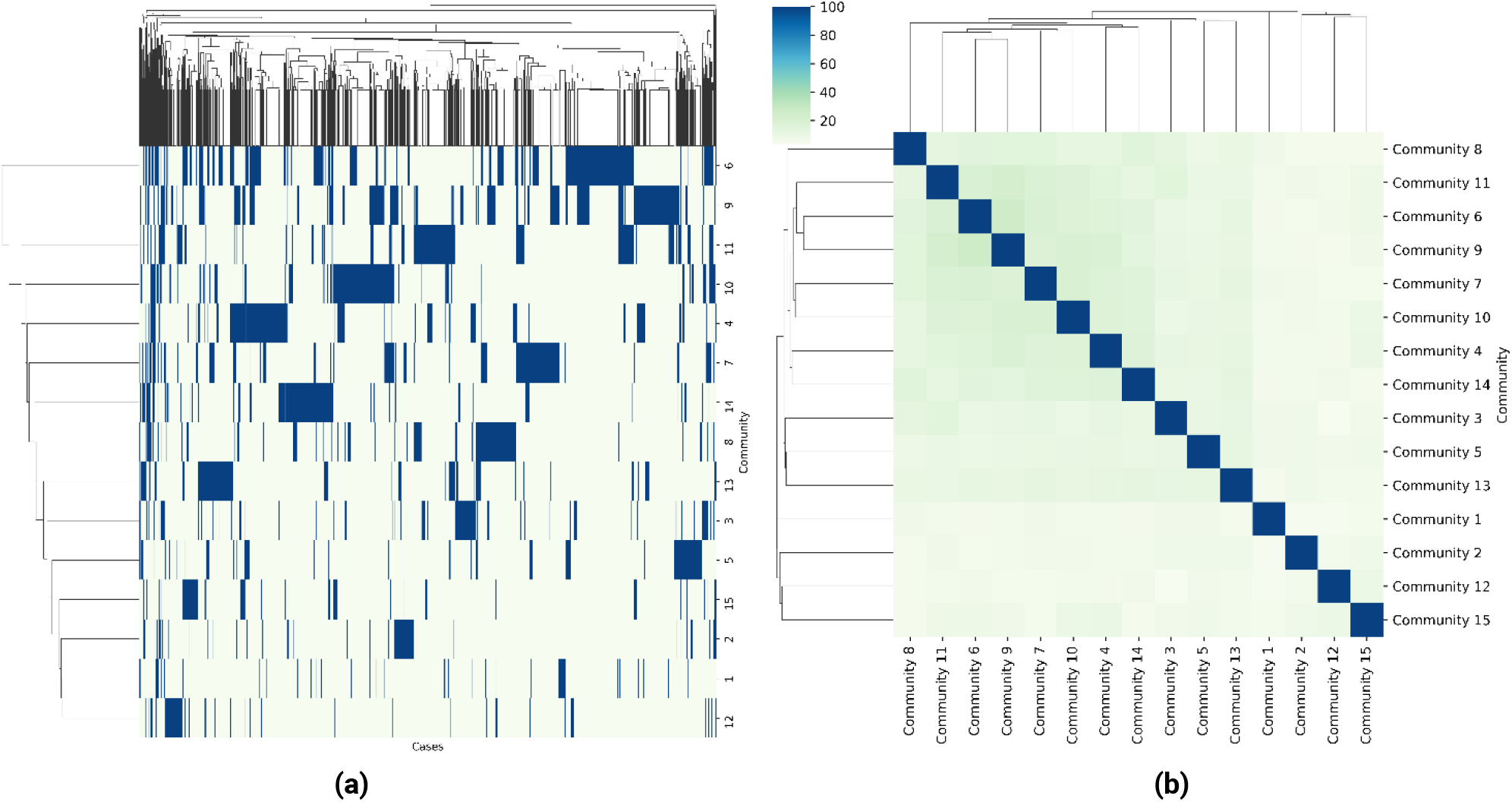
**(a)** Clustered heatmap showing the similarity of the 15 communities based on their respective cohorts of associated Pain Questionnaire patients. (**b)** Clustered heatmap showing the overlap of Pain Questionnaire patients associated with the 15 communities identified in the ME/CFS disease architecture.

**Figure 7:**
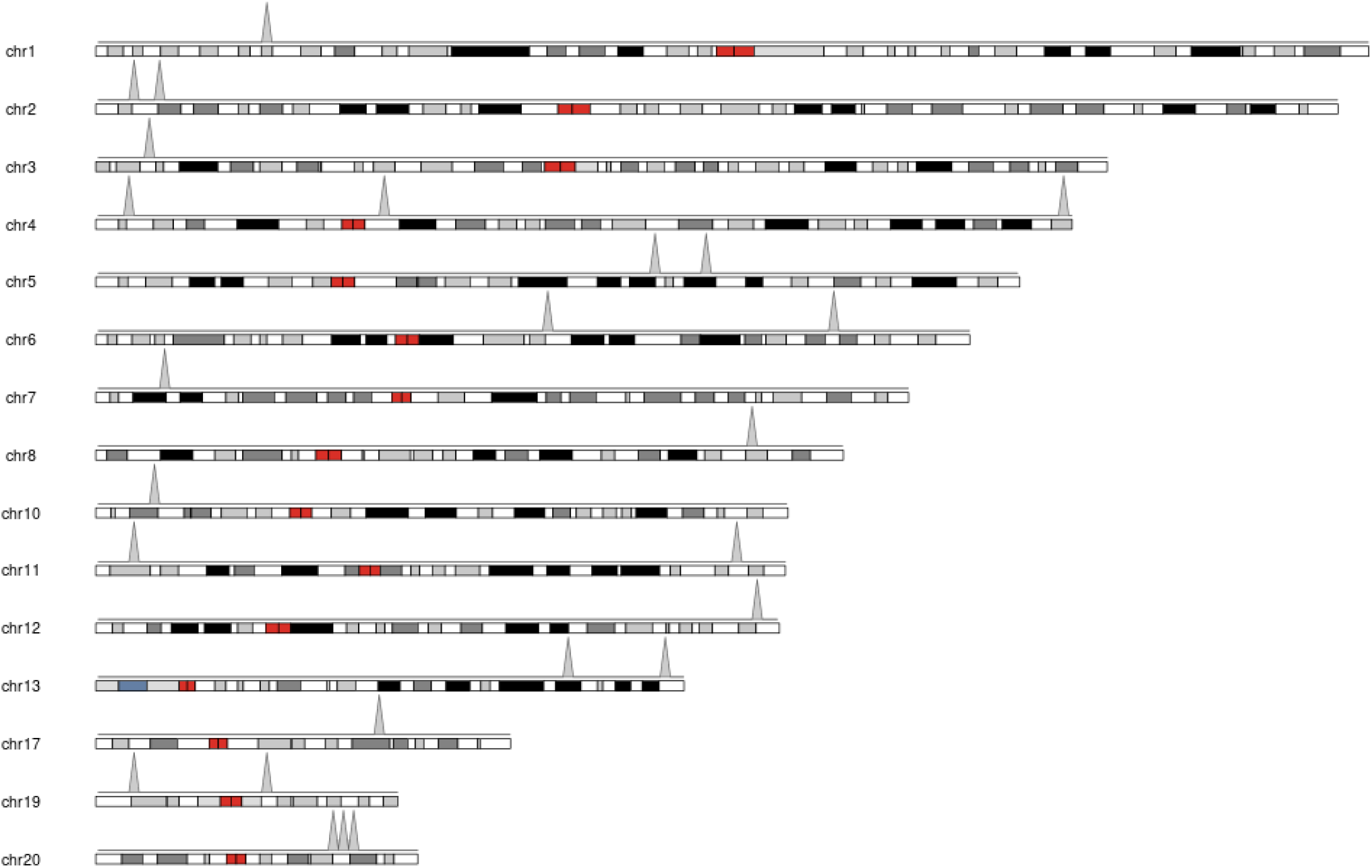
Autosomal locations for 25 critical disease associated SNPs identified in the Pain Questionnaire cohort.

**Figure 8:**
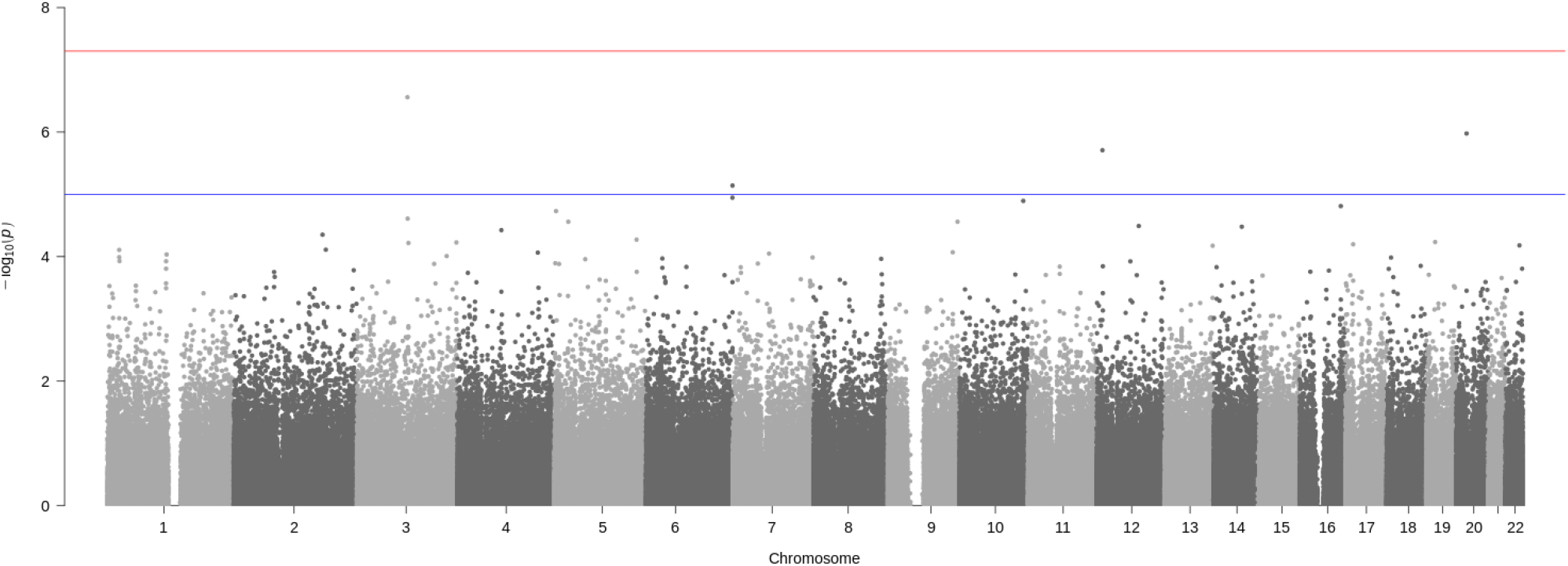
Manhattan plot generated using PLINK of genome-wide *p*-values of association for the disjoint Verbal Interview cohort (n = 5,140 where cases=1,273 and controls=4,137). The horizontal blue and red lines represent the genome-wide significance values of *p*<1 × 10^−5^ and *p*<5 × 10^−8^ respectively.

**Figure 9:**
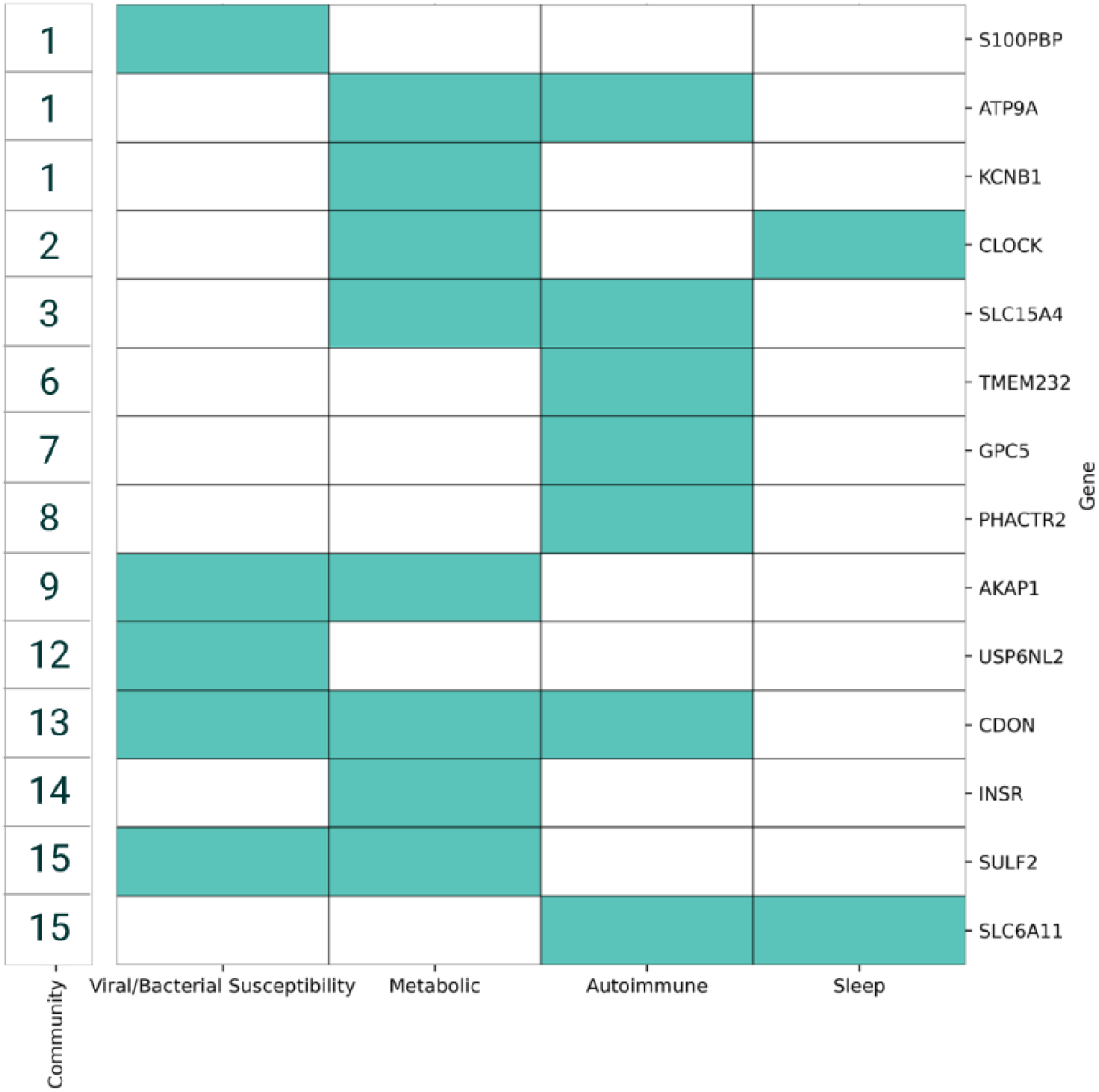
Biological pathways and processes known to be associated with the genes identified by the Pain Questionnaire study. Each border color represents a different patient community.

**Figure 10:**
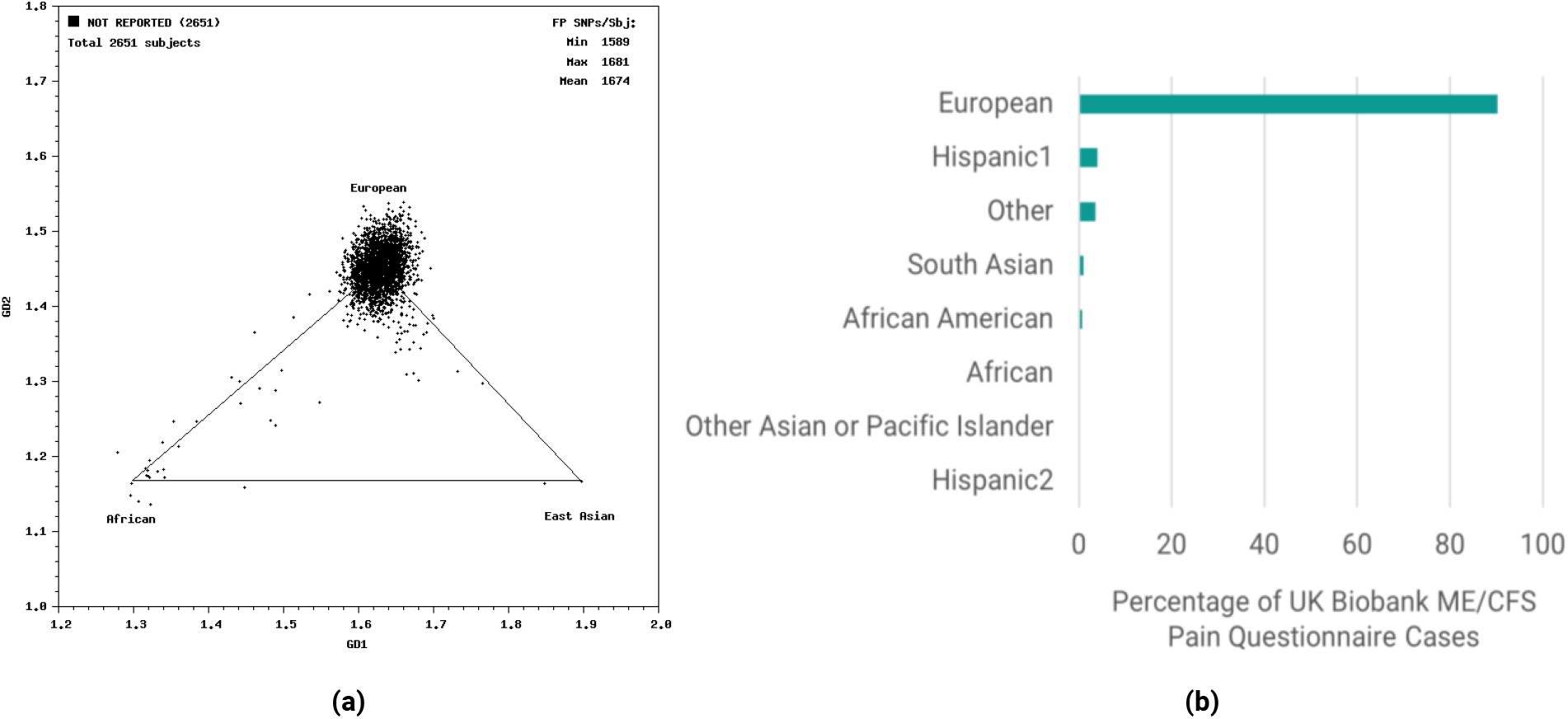
(a) Ancestry inference plot generated by GRAF-pop and (b) the ancestry distribution of ME/CFS case population (n=2,651 cases) generated from UK-Biobank using self-reported diagnosis in Pain Questionnaire before quality control shows very strong bias for European ancestry (>90%).

**Figure 11:**
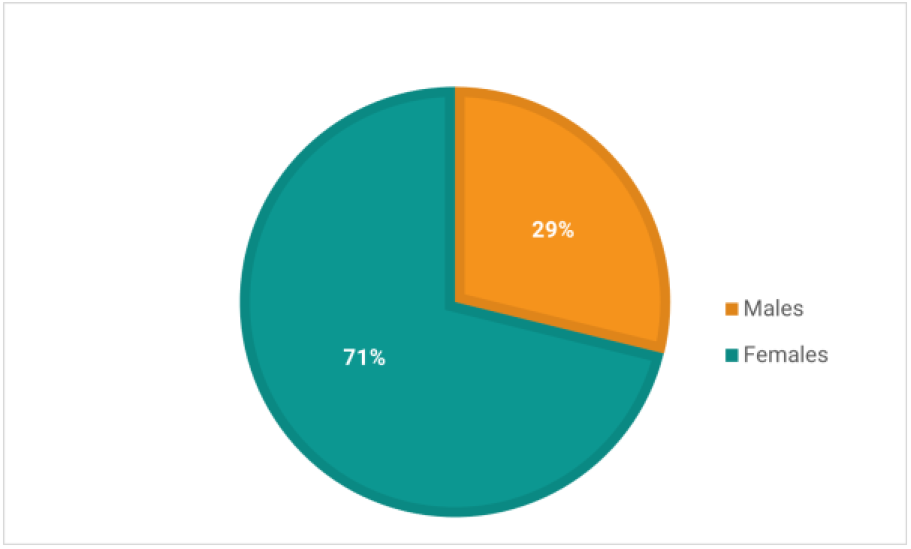
The proportion of females in the Pain Questionnaire case population was substantially higher (∼71%) than males (29%).

**Figure 12:**
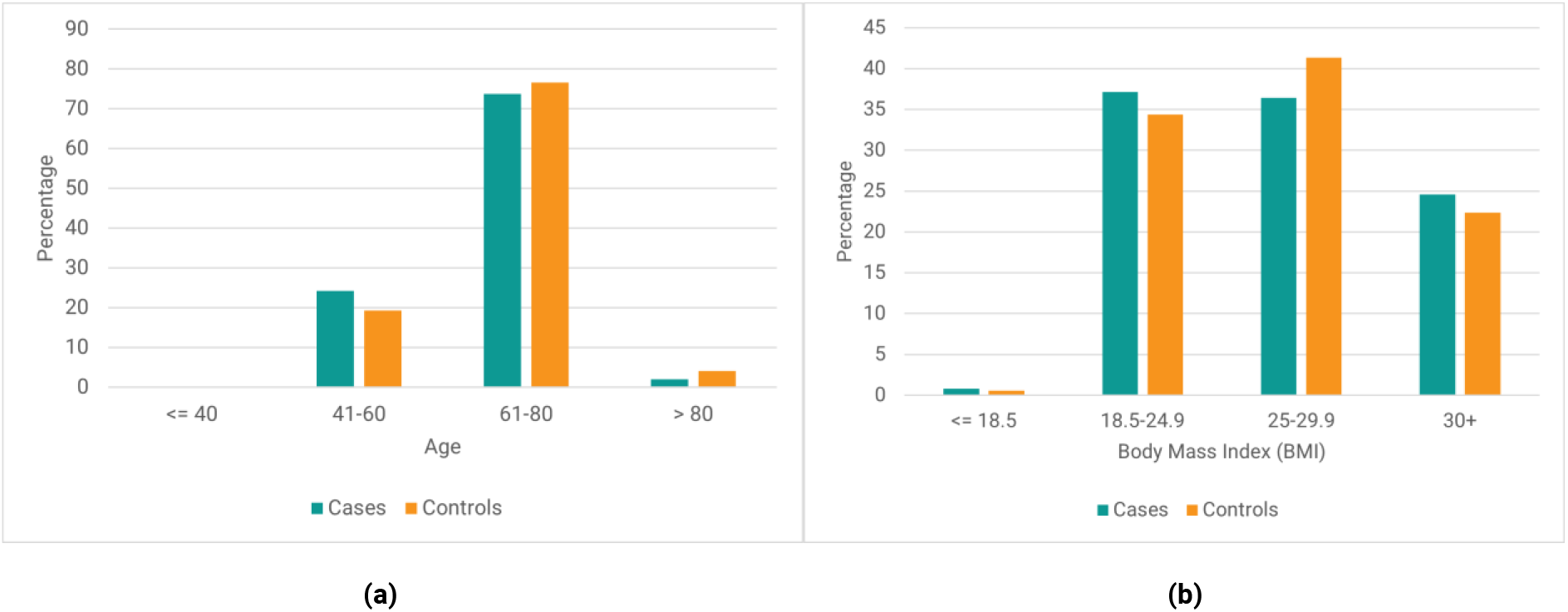
Distribution of (a) age and (b) BMI of cases vs controls in the Pain Questionnaire cohort.

Data about individuals’ diagnosis with autoimmune disease (including multiple sclerosis and fibromyalgia), self-reported emotional or physical stress, and exposure to potential viral triggers (such as EBV seropositivity) were used to compare ME/CFS cases against the remainder of the UK Biobank to identify any significant differences between the two cohorts that could be associated with ME/CFS onset (see Figure 2).

After quality control (see Genotype Quality Control in Supplementary Data), the Pain Questionnaire dataset was comprised of 2,382 ME/CFS cases, 4,764 controls and 519,337 SNPs on autosomal chromosomes. Approximately 71% of cases (n=1,695) were women (**Error! Reference source not found**.Figure 11 in Supplementary Data) vs the UK Biobank distribution of 54.4%. The age and body-mass index (BMI) distributions of cases and controls were similar (Figure 12b in Supplementary Data).

### Methods

We applied the PrecisionLife platform to the various ME/CFS case-control datasets to identify combinations of SNP genotypes that when observed together in a sample were strongly associated with the development of ME/CFS. The PrecisionLife platform uses a unique data analytics framework that enables efficient combinatorial analysis of large, multi-dimensional participant datasets. Navigating this data space allows for the identification of combinations of features that are significantly associated with groups of cases in a case-control dataset. The PrecisionLife combinatorial analysis is hypothesis free, involving a four-stage mining, validation, evaluation and annotation process.

The PrecisionLife platform identifies combinations of feature states in ‘layers’ of increasing combinatorial complexity, i.e., singletons, pairs, triplets etc. A feature could for example be a SNP, and a feature state would consist of the SNP’s base index and its genotype, which would typically be encoded ordinally as {0, 1, 2} for homozygous major allele, heterozygous minor allele, homozygous minor allele respectively. The platform has considerably more flexibility of representation (including alternate genotype encodings, extended genetic models, polyploidy and quantitative values) if required by the feature or dataset being analyzed.

In the mining phase, combinations of feature states that are overrepresented (using a Z-score or Fisher’s Exact test) in cases are identified and validated (Table 1). Multiple feature states are combined iteratively until no additional features can be added that will improve the score. Combinations of feature states that have high odds ratios, low *p*-values (*p* < 0.05) and high prevalence (>5%) in cases are prioritized. The mining process is repeated across up to 1,000 cycles of fully randomized permutation of the case:control labels of all individuals in the dataset, keeping the same parameters and case-control ratio.

In the validation phase, all combinations generated by the original mining run and each of the random permutation iterations of the dataset are compared. These combinations are validated using network properties such as minimum prevalence (number of cases represented, in this case >5%) as the null hypothesis when compared with the combinations generated by the random permutations. Combinations that appear in the random permutations above a specified FDR threshold (Benjamini-Hochberg FDR of 0.05) after multiple testing correction^23^ are considered to be random and eliminated. Combinations passing these tests are reported as validated disease signatures.

The validated disease signatures are then evaluated. The features (which in this case only consisted of SNPs due to the limited available dataset) shared by multiple disease signatures (known as ‘critical’ SNPs) are identified. Critical SNPs, which can be thought of as the canonical features of a cluster comprised of overlapping disease signatures, are then scored using a Random Forest (RF) algorithm in a 5-fold cross-validation framework to evaluate the accuracy with which they predict the observed case-control split in a dataset (minimizing Gini impurity or the probability of misclassification).

We use RF scores in similar ways to rank critical SNPs and by association the genes to which they map via the process described in the Functional Genomics Annotation section below. Disease signatures comprising high RF scoring critical SNPs (and their genes) are then mapped to the cases in which they were found, and additional clinical data (such as blood biochemistry data, comorbidity ICD-10 codes and medication history) is used to generate a patient profile for each combinatorial disease signature.

Finally, a merged network (disease architecture) view is generated by clustering all validated disease signatures based on their co-occurrence in patients in the dataset, and annotation of the validated SNPs, genes, and the druggability of targets is performed using a semantic knowledge graph (see Functional Genomics Annotation section).

For these studies, the PrecisionLife platform generated statistically significant ME/CFS associated signatures containing up to five SNPs for each cohort. Each ME/CFS dataset analysis took around 7 days (168) hours to complete, running on a server with 64 CPU cores and 4x Nvidia GPUs.

### Replication and Validation

No similarly sized ME/CFS cohort is currently available for use as an independent replication study cohort. We therefore used two alternate approaches to validate the results from the Pain Questionnaire study.

In the first approach, we performed the 1,000 random permutation tests on each combinatorial disease signature by randomly shuffling cases and controls in the Pain Questionnaire dataset and calculating a permutation test score (P1000) for all observed SNP combinations across the full range of combinatorial order. The P1000 score of a combinatorial disease signature indicates the frequency of detection of similarly associated combinatorial features in the 1,000 randomized permutations, as measured by odds ratio and number of cases possessing the feature. Any feature where P1000 is less than 50 (i.e., 5%) is considered significant.

In a second approach, we generated three new case populations from UK Biobank for fatigue-associated conditions and compared the results from these with the Pain Questionnaire cohort. These included a new CFS case population (Data-Field 20002, Coding 1482) reported during Verbal Interview, a post-viral syndrome (ICD10: G93.3) case population based on Hospital Episode Statistics, and a fibromyalgia (ICD10: M79.7) case population based on Hospital Episode Statistics.

The Verbal Interview case population comprised of 2,270 individuals whose data had been analyzed in a recent GWAS study^24^. As the Verbal Interview cohort had 735 individuals in common with the Pain Questionnaire cohort (Figure 17 in Supplementary Data), these overlapping cases were removed to create a disjoint Verbal Interview dataset (cases = 1,273 and controls = 4,137 after QC) of European ancestry with gender (and ancestry) matched controls. Similarly, disjoint post-viral syndrome dataset (cases = 510 and controls = 4,763) and fibromyalgia dataset (cases = 1,409 and controls = 4,762 after QC) of European ancestry were generated.

**Figure 13:**
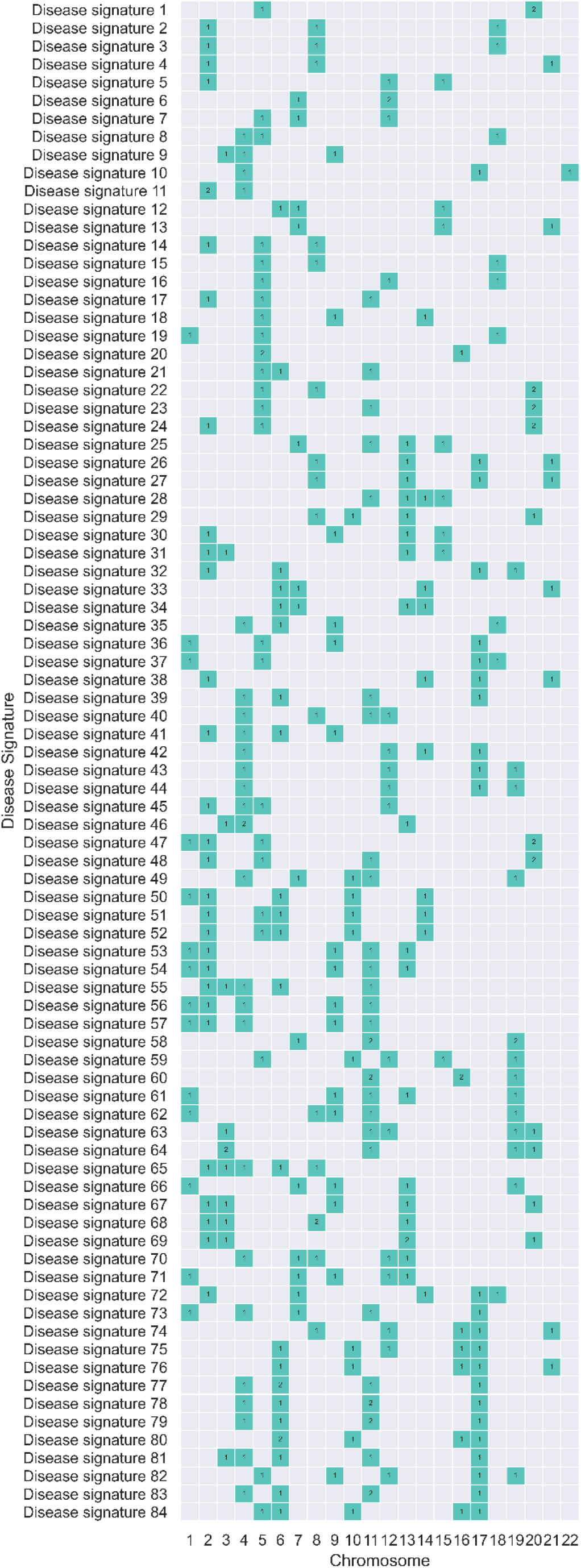
Distribution of chromosomal location of SNPs associated with 84 disease signatures identified in the Pain Questionnaire study. None of the SNPs identified in the disease signatures were observed to be in linkage disequilibrium (LD).

**Figure 14:**
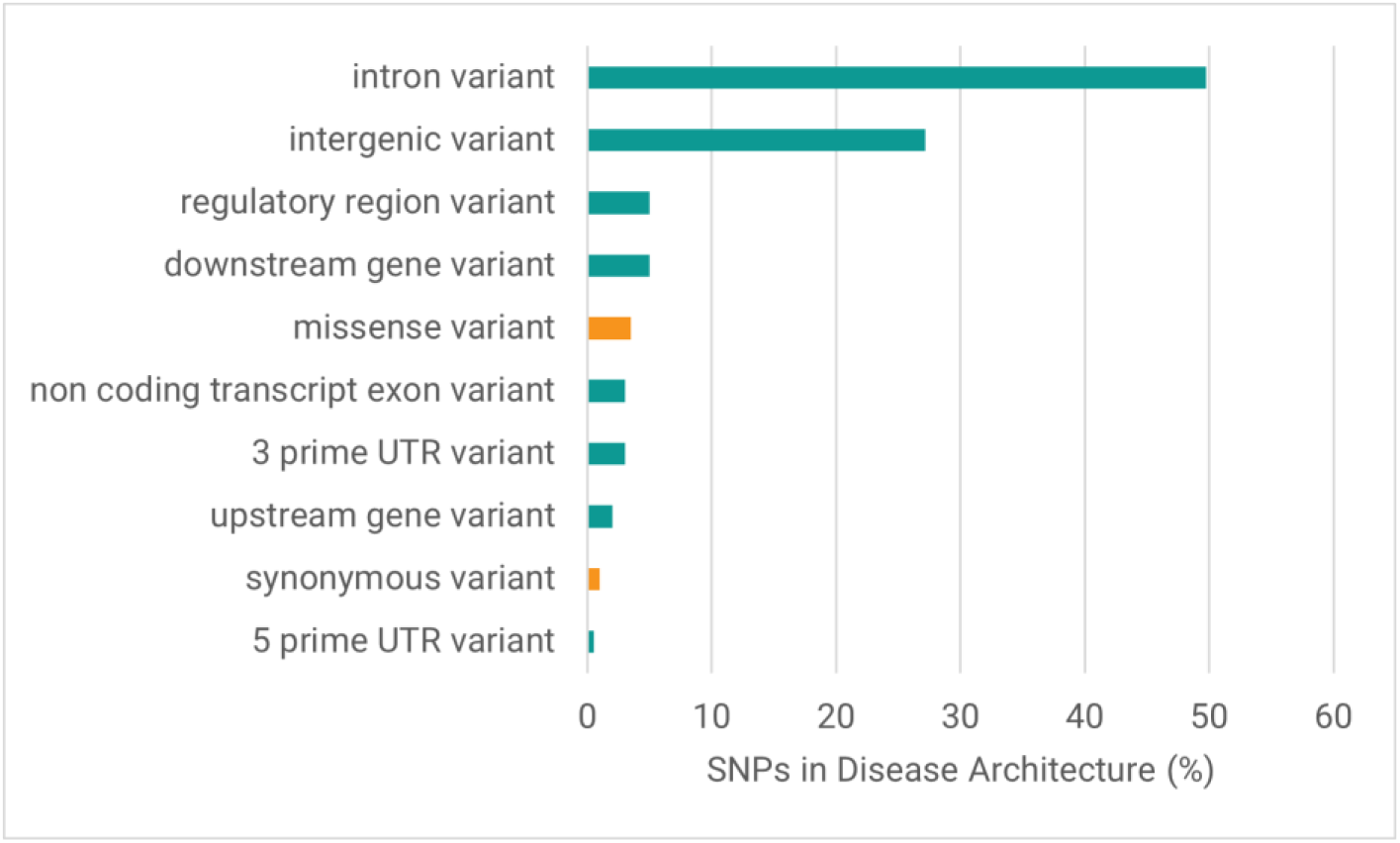
Distribution of variant consequences (most severe predicted by Ensembl VEP) of critical SNPs identified in the Pain Questionnaire study. More than 95% of SNPs were non-coding variants (shown in green) and <5% were coding variants (shown in orange).

**Figure 15:**
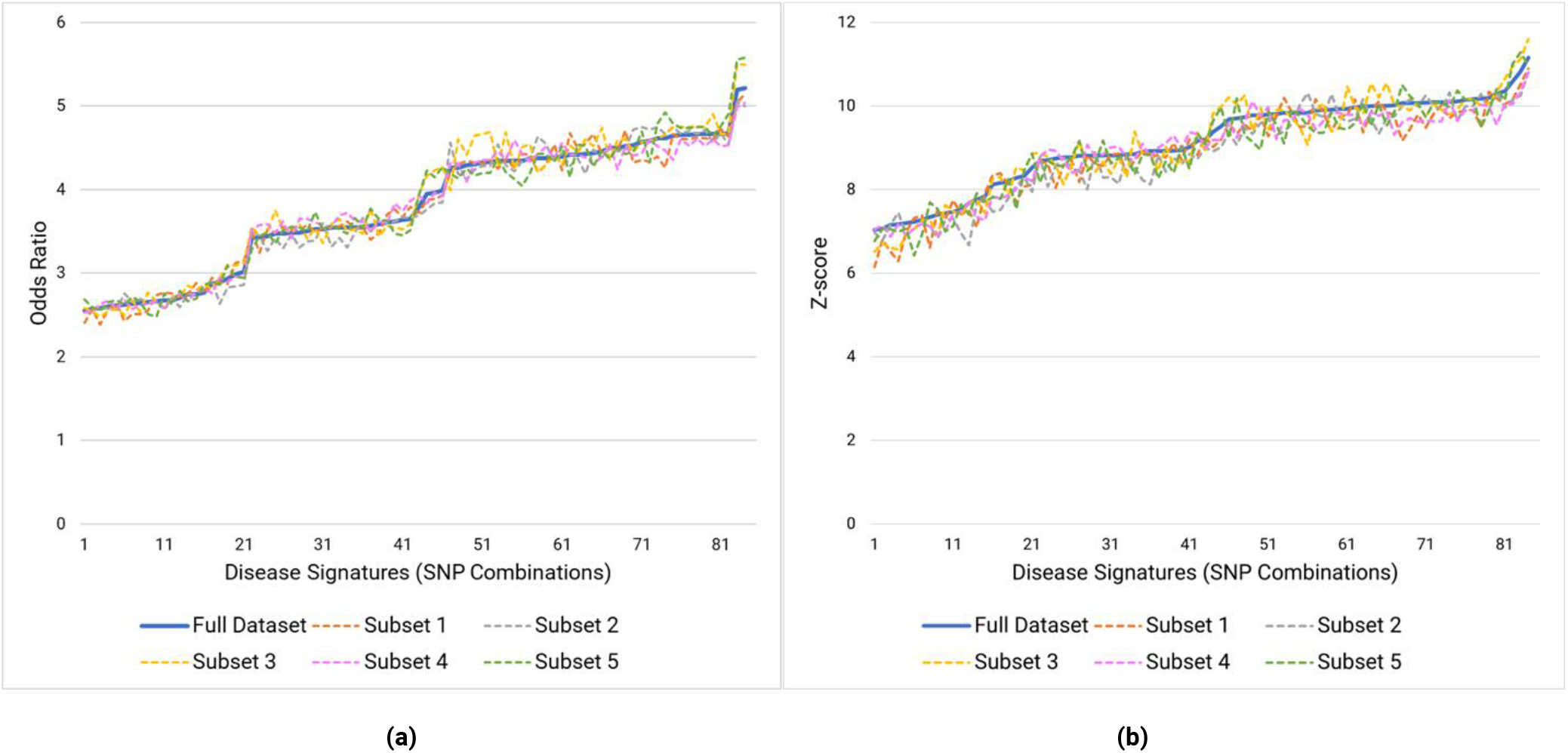
Comparison of (a) odds ratios and (b) Z-scores of the full Pain Questionnaire dataset with five sub-cohort splits containing 90% of the same cases as the full cohort and the same set of controls.

**Figure 16:**
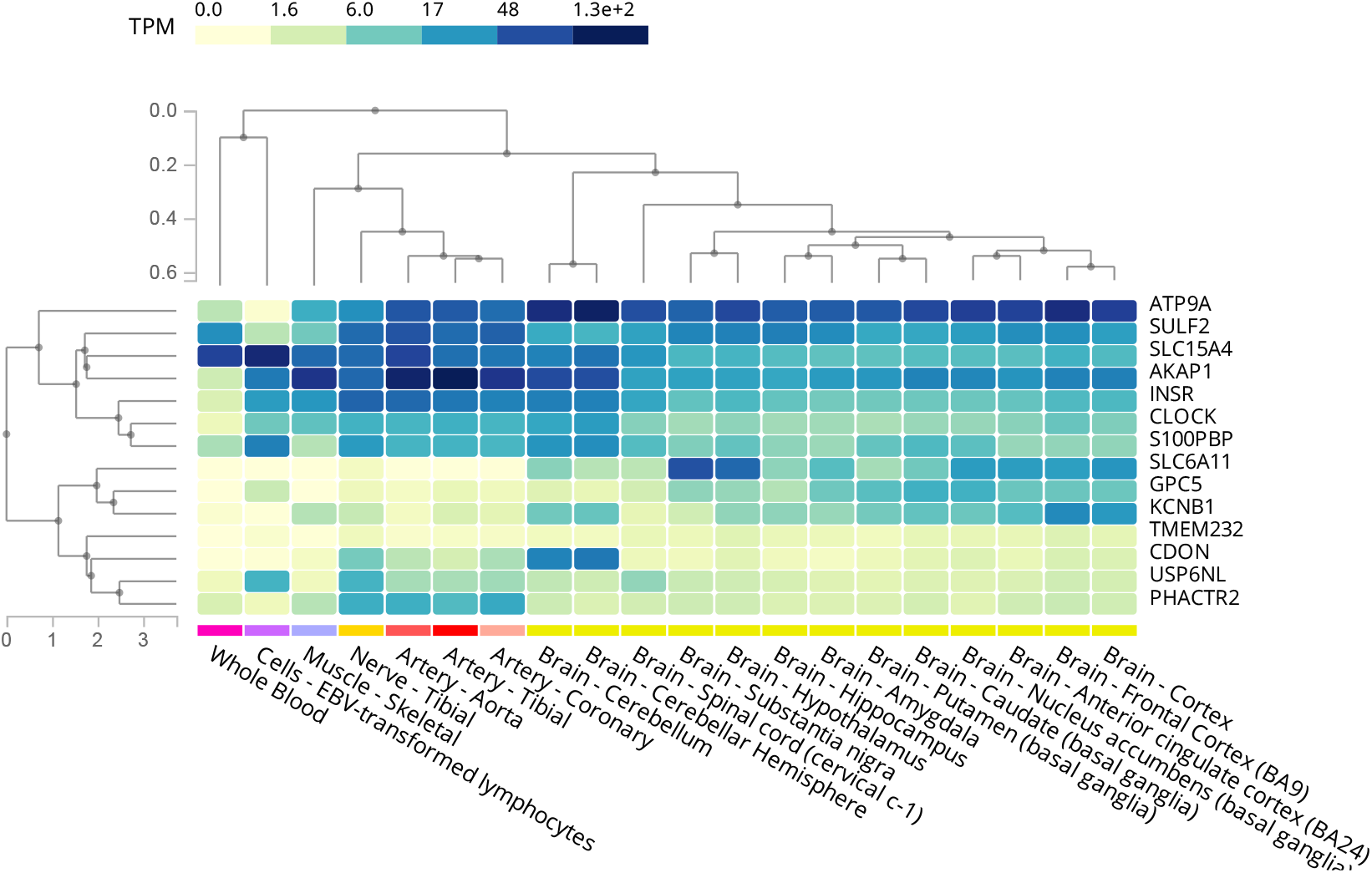
Clustered heatmap showing tissue expression profiles (GTEx) for 14 genes identified in the Pain Questionnaire study.

**Figure 17:**
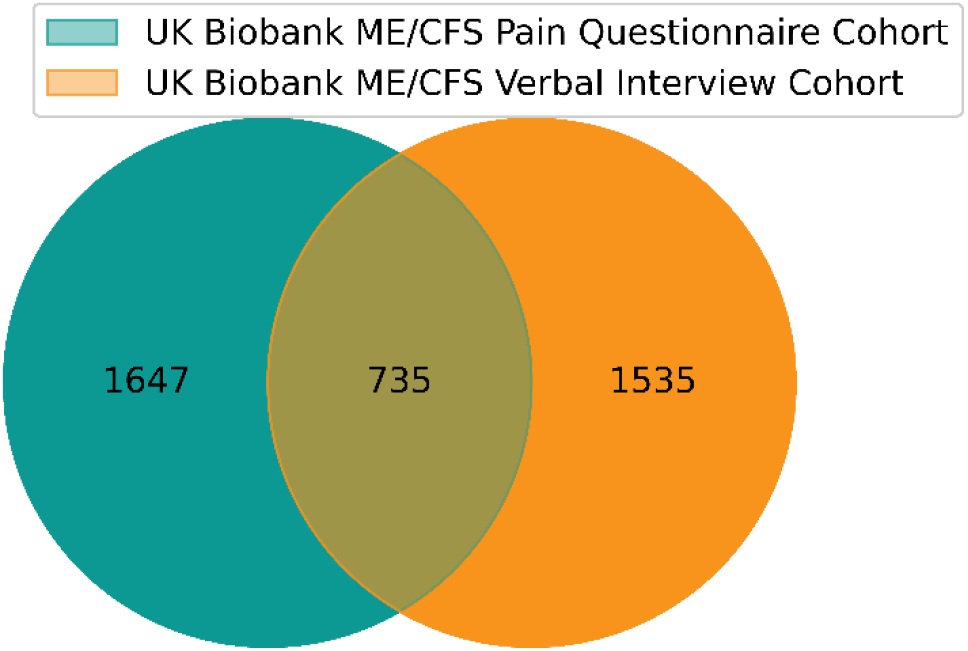
Case overlap between two UK Biobank ME/CFS cohorts (Pain Questionnaire and Verbal Interview)

The three disjoint datasets (Verbal Interview CFS, post-viral syndrome and fibromyalgia) were also analyzed through the PrecisionLife platform to investigate the extent to which the results generated from the original Pain Questionnaire cohort could be replicated in these cohorts.

Two issues contribute to limit the degree of overlap that can be expected in a fully independent analysis of the disjoint cohorts. Firstly, as the combinatorial search space is vast, the sampling of that space is likely to be materially incomplete, which will contribute to a potential high rate of false negatives, i.e. true associations that were not tested or reported due to random sampling bias. A separate more systematic sampling of the space will be run in future studies, but this method was not available for this study. Secondly, within the two ME/CFS clinical diagnoses, the assignment by a GP of either a CFS or ME/CFS or post-viral syndrome diagnosis is highly variable and cannot be relied upon to distinguish these case populations in a clinically meaningful manner.

Due largely to the first sampling issue, it is likely that these three studies will differ significantly from the Pain Questionnaire study in the reported similarity of their genetic associations and clinical characteristics.

We therefore limited the search space for the analysis of the three disjoint cohorts by testing only combinations involving the 199 SNPs identified in the Pain Questionnaire cohort. Limiting the search space to combinations involving these critical SNPs enables us to assess the level of replication of the ME/CFS genetic signal in the second dataset by eliminating the unavoidable sampling bias arising from differences in the heterogeneous patient populations exacerbated by the small numbers of case available in the huge search space.

### Functional Genomics Annotation

We mapped all SNPs identified in the disease signatures using an annotation cascade process to the human reference genome (GRCh37)^25^ to give the best estimate of the gene(s) likely to be associated with the SNP. Disease-associated SNPs that lie within coding regions of gene(s) were assigned directly to the corresponding gene(s). Remaining SNPs that lie within 2kb upstream or 0.5kb downstream of any gene(s) were mapped to the closest gene(s) within this region. The potentially causality and druggability of these genes were evaluated in later steps.

We investigated additional gene assignments for the identified SNPs using publicly available eQTL^26^ and/or chromatin interaction data^27^ (see Supplementary Table 11). Genes with at least one cis-eQTL SNP at a false discovery rate (FDR) of ≤0.05, with expression differences of that gene in single brain tissues or whole blood, were reported^26^.

Additionally, promoter capture Hi-C (pcHi-C) interactions that were significantly associated in brain tissues and blood cells by Jung et. al ^27^ were used to generate gene assignments. Due to the uncertainty about the relevant cells and tissues affected in ME/CFS etiology, genes assigned by either eQTL or chromatin interaction data were not specifically prioritized for further analysis (as they might be in other studies) to avoid capturing any spurious associations from non-trait-related tissues^26^. Genes that could be additionally mapped using only eQTL or HiC data from the 25 critical SNPs were however observed and reported in Supplementary Table 11, although these were not further evaluated. The direction of association of any eQTLs associated with the disease phenotype was however noted.

Critical SNPs (see Methods section) were assigned an RF score, describing how well the SNP genotype combinations predict the observed case-control split. We used these scores to rank the critical SNPs to reflect the relative importance of the SNP and its combinations. The genes assigned to the critical SNPs were prioritized on the basis of the cumulative sum of their associated SNP scores to identify the most clinically relevant targets, as the critical SNPs are those observed to have markedly higher association with the disease.

We used a semantic knowledge graph derived from over 50 public and private data sources to annotate the prioritized genes (see Supplementary Table 12). This included information from a variety of data sources including basic genomic context, tissue expression, chemical tractability, biological function and associated scientific literature. We tested each of the genes identified against the 5Rs criteria^28^ of early drug discovery, to form and validate hypotheses for their mechanism of action and impact on the disease phenotype.

### Patient Stratification

The output disease signatures generated by the PrecisionLife platform contain metadata including the indices of all the cases (and controls) in which they were found. The available phenotypic and clinical data for the relevant cases were used to evaluate patient profiles associated with each of the disease signatures. This was based on the observed enrichment of an attribute or phenotype for a particular group of patients (for example association with a prioritized gene) compared against the entire case population. Statistical significance was calculated using the two proportions Z-test for categorical variables such as gender and co-morbidities, whereas we used the Mann-Whitney U test for continuous variables such as measurements of metabolic biomarkers. *p*-values corrected for multiple-testing using the Benjamini-Hochberg method to control the FDR were also reported.

## Results

### UK Biobank ME/CFS (Pain Questionnaire) Cohort Characteristics

We identified significant differences in a variety of covariates (listed in Cohort Analysis section in Supplementary Data) between the ME/CFS case population, and the control group, and the remaining individuals in UK Biobank (Figure 2). The figure shows the percentage of individuals in each group who are positive for each covariate. To test for significance, we calculated 95% confidence intervals using bootstrapping (sampling with replacement) for 1,000 iterations.

The greatest difference between the ME/CFS population in this study and the remainder of the UK Biobank was the significantly higher proportion of individuals reporting mental distress and stressful events such as illness, injury, and bereavement. Individuals with ME/CFS in this study were also slightly more likely to present with at least one autoimmune disease, with the greatest co-association with other myalgia and fatigue-associated conditions like multiple sclerosis and fibromyalgia. It is however impossible to rule out a level of misdiagnosis in these complex conditions.

### Combinatorial Analysis

The ME/CFS Pain Questionnaire cohort (2,382 cases, 4,764 controls) was used to perform a standard GWAS case-control association analysis using PLINK^29^. No SNPs were reported to be significant below a genome-wide significance threshold of *p*<5 × 10^−8^ (Figure 3).

Combinatorial SNP analysis performed using the PrecisionLife platform on the same Pain Questionnaire dataset generated 84 statistically validated combinations of 199 SNPs that together are strongly associated with ME/CFS diagnosis (Table 2, Table 10 in Supplementary Data). None of the SNPs identified were observed to be in linkage disequilibrium (LD) (Figure 13 in Supplementary Data). 192 SNPs identified in the disease signatures were in non-coding regions of the genome and 9 (7 missense and 2 synonymous) SNPs were identified in the coding regions (Figure 14 in Supplementary Data).

**Table 10:**
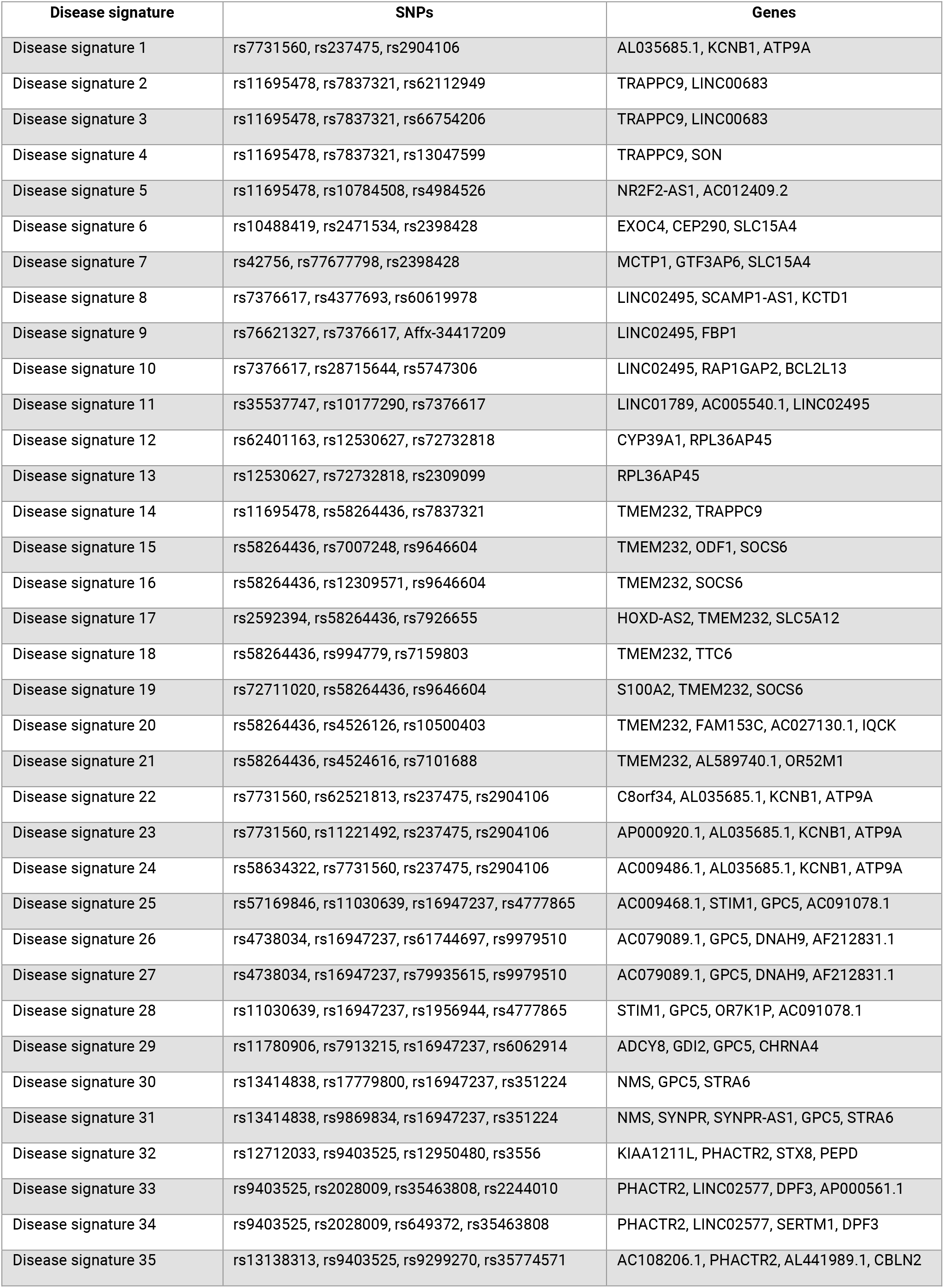
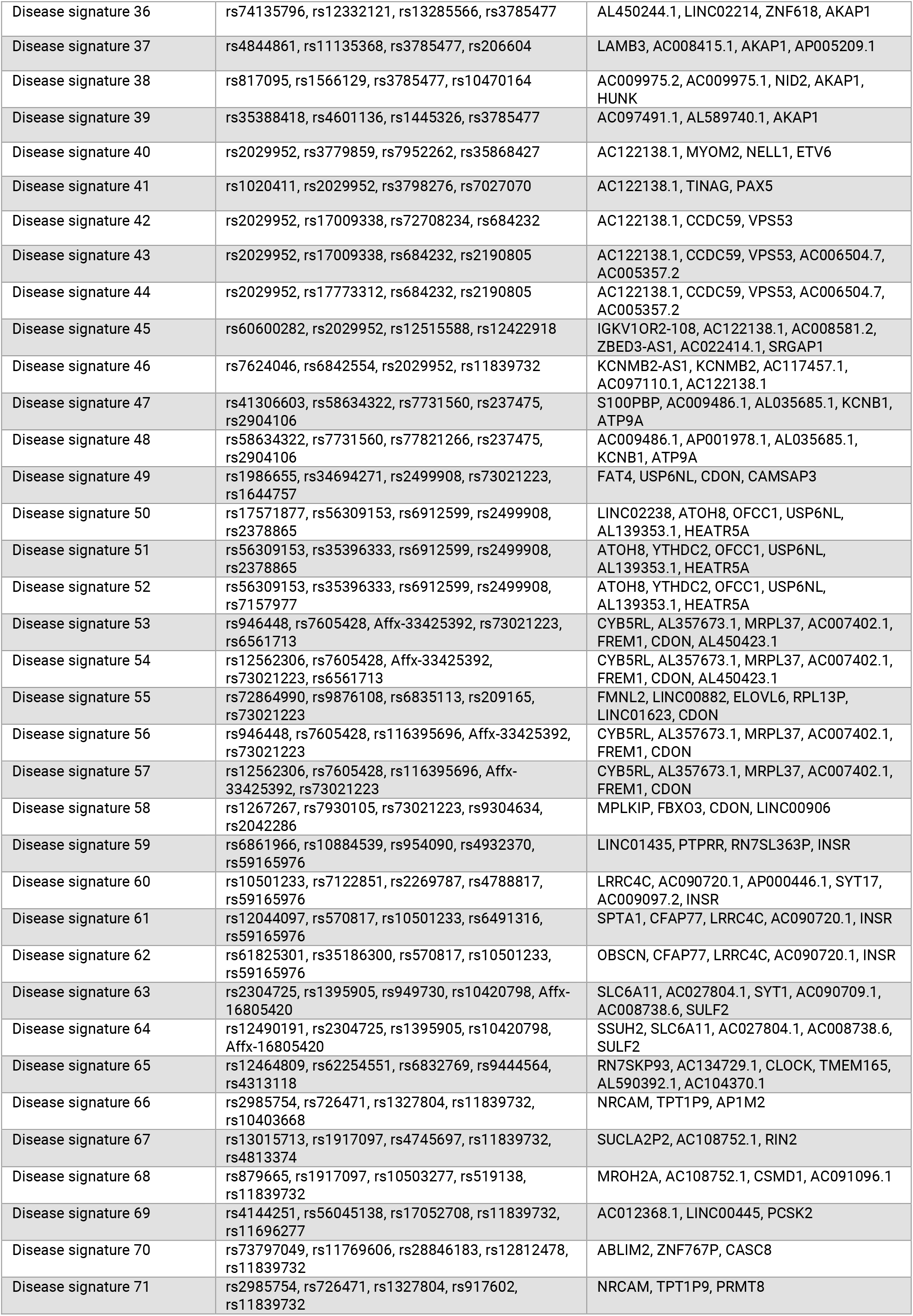
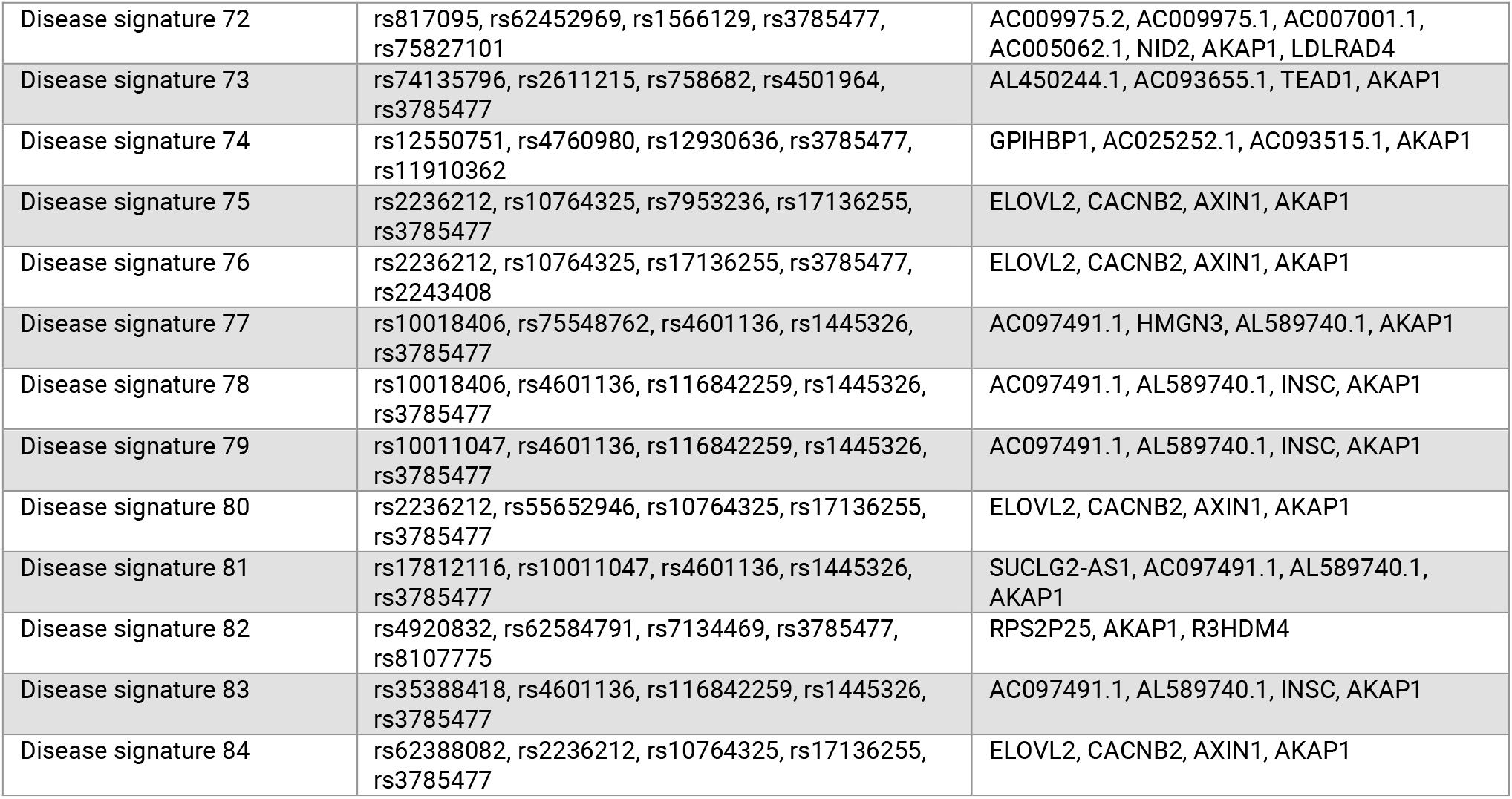
List of 84 disease signatures with its constituent SNPs and mapped genes identified in the ME/CFS Pain Questionnaire study.

All SNPs were found in combinations with 3 or more SNPs, and so would not have been found using standard GWAS analysis methods (Figure 4a, Table 10 in Supplementary Data). No single SNPs or SNP pairs were reported as significant by the method.

As a negative validation check, runs using the same mining and validation parameters as used above were performed on 7,146 random samples, comprising 2,382 UK Biobank randomly sampled participants as ‘cases’ compared against 4,764 randomly sampled ‘controls’. This analysis yielded no significant results.

These 84 combinatorial disease signatures all had P1000 values of 0, indicating they were not detected in any of the 1,000 random permutation runs, and are therefore very unlikely to result from random chance (Figure 4b). The odds ratios of the SNP combinations were found to be around 3.7 on average (Figure 4b).

Table 3 represents an example of a disease signature identified in this analysis containing five SNPs that were mapped to five genes.

### Patient Stratification

The disease architecture (Figure 5) generated by clustering^30^ the SNPs in the disease signatures on the basis of patients in which they co-occur reveals the genetic heterogeneity of the ME/CFS Pain Questionnaire patient population, providing useful insights into patient stratification. These clusters (‘communities’) represent patient subgroups that (by definition) have shared disease etiology, and are therefore likely to share disease phenotypes, including severity, progression rate, clinical presentation, and, ultimately, therapy response.

There are 15 distinct communities of SNPs shown in the ME/CFS Pain Questionnaire disease architecture (Table 4 Table 4). These share very low (<20%) patient overlap with each other (Figure 6), indicating they are distinct patient subgroups with different genetic drivers underlying their disease.

The analysis identified 25 critical disease associated SNPs (see the Methods and Functional Genomics Annotation sections) which are identified in multiple disease signatures. These critical SNPs (Figure 7) were mapped to 14 protein coding genes strongly associated with the ME/CFS Pain Questionnaire case population (Table 5). Investigation of the function and mechanisms of action of these genes (and encoded proteins) revealed associations with one or more of five disease mechanisms that have been associated with ME/CFS development – viral/bacterial susceptibility, autoimmune development, metabolic dysfunction, vulnerability to stress, and sleep disturbance.

Enrichment analysis of available phenotypic and clinical data for the ME/CFS patients was used to generate additional insights into the clinical characteristics of each SNP community and prioritized gene. Statistical significance was calculated using the two proportions Z-test for categorical variables and the Mann-Whitney U test for continuous variables.

This analysis revealed 11 genes from 6 different patient communities with a level of enrichment with a particular phenotypic or clinical feature, such as increased incidence of clinical diagnosis of fibromyalgia or increased phenylalanine levels in plasma (Table 5), when compared against the rest of the case population. These associations, however, did not reach statistical significance (*p*<0.05) after multiple testing correction (Table 11 and Table 12 in Supplementary Data).

**Table 11:** Frequency distribution of categorical phenotypic features in cases associated with gene(s) compared to all cases and controls in the cohort. *p*-values were calculated to assess the association of each feature using two-sided Fisher’s exact tests.

| Phenotypic Feature | Gene(s) | Gene case count with feature | Gene Case count | All case count with feature | Total case count | All controls count with feature | Total control count | *p*-value (compared to cases) before multiple testing | Adjusted *p*-value (compared to cases) | *p*-value (compared to controls) before multiple testing | Adjusted *p*-value (compared to controls) |
| --- | --- | --- | --- | --- | --- | --- | --- | --- | --- | --- | --- |
| Illness injury bereavement stress in last 2 years (2019+) | *SLC6A11*<br>*SULF2* | 4 | 170 | 12 | 2,382 | 1 | 4,764 | 0.003 | 0.177 | $6.00 \times 10^{-21}$ | $7.61 \times 10^{-21}$ |
| Illness injury bereavement stress in last 2 years (2014+) | *CDON* | 48 | 328 | 249 | 2,382 | 3 | 4,764 | 0.023 | 0.861 | $6.36 \times 10^{-145}$ | $1.19 \times 10^{-144}$ |
| Fibromyalgia (ICD-10: M79.7) | *CLOCK* | 13 | 153 | 108 | 2,382 | 0 | 4,764 | 0.026 | 0.969 | $2.92 \times 10^{-90}$ | $1.87 \times 10^{-89}$ |
| Other soft tissue disorders (ICD-10: M79) | *CLOCK* | 22 | 153 | 224 | 2,382 | 0 | 4,764 | 0.044 | 0.969 | $1.15 \times 10^{-151}$ | $1.04 \times 10^{-149}$ |
| Thyroiditis (ICD10-10: E06) | *PHACTR2* | 3 | 343 | 5 | 2,382 | 0 | 4,764 | 0.033 | 0.969 | $1.07 \times 10^{-10}$ | $1.92 \times 10^{-10}$ |
| Males (Genetic sex) | *S100PBP* | 48 | 125 | 687 | 2,382 | 11 | 4,764 | 0.022 | 0.078 | 0 | 0 |
| Males (Genetic sex) | *ATP9A*<br>*KCNB1* | 55 | 144 | 687 | 2,382 | 31 | 4,764 | 0.016 | 0.078 | $7.29 \times 10^{-251}$ | $3.28 \times 10^{-250}$ |

**Table 12:** Frequency distribution of quantitative phenotypic features in cases associated with gene(s) compared to all cases and controls in the cohort. *p*-values were calculated to assess the association of each feature using two-sided Mann-Whitney U tests.

| Phenotypic Feature | Gene(s) | Gene case count with feature data | Max, Mean & Min value of feature for cases with gene assoc. | All case count with feature data | Max, Mean & Min value of feature for all cases | Control count with gene assoc. and feature data | Max, Mean & Min value of feature for controls | *p*-value (compared to cases) before multiple testing | Adjusted *p*-value (compared to cases) after multiple testing |
| --- | --- | --- | --- | --- | --- | --- | --- | --- | --- |
| Phenylalanine levels in plasma | *SLC6A11*<br>*SULF2* | 41 | 0.07<br>0.047<br>0.032 | 595 | 0.08<br>0.044<br>0.017 | 0 | N/A | 0.022 | 0.988 |

**Table 13:** Frequency distribution of quantitative phenotypic features in cases associated with communities compared to all cases and controls in the cohort. *p*-values were calculated to assess the association of each feature using two-sided Mann-Whitney U tests.

| Phenotypic Feature | Community | Gene case count with feature data | Max, Mean & Min value of feature for cases with gene assoc. | All case count with feature data | Max, Mean & Min value of feature for all cases | Control count with gene assoc. and feature data | Max, Mean & Min value of feature for controls | *p*-value (compared to cases) before multiple testing | Adjusted *p*-value (compared to cases) after multiple testing |
| --- | --- | --- | --- | --- | --- | --- | --- | --- | --- |
| Lactate levels in plasma | 3 | 61 | 1.81<br>3.41<br>6.81 | 595 | 1.41<br>3.69<br>9.64 | 0 | N/A | 0.028 | 0.992 |
| Lactate levels in plasma | 14 | 100 | 1.66<br>3.92<br>7.13 | 595 | 1.41<br>3.69<br>9.64 | 0 | N/A | 0.017 | 0.992 |

**Table 14:**
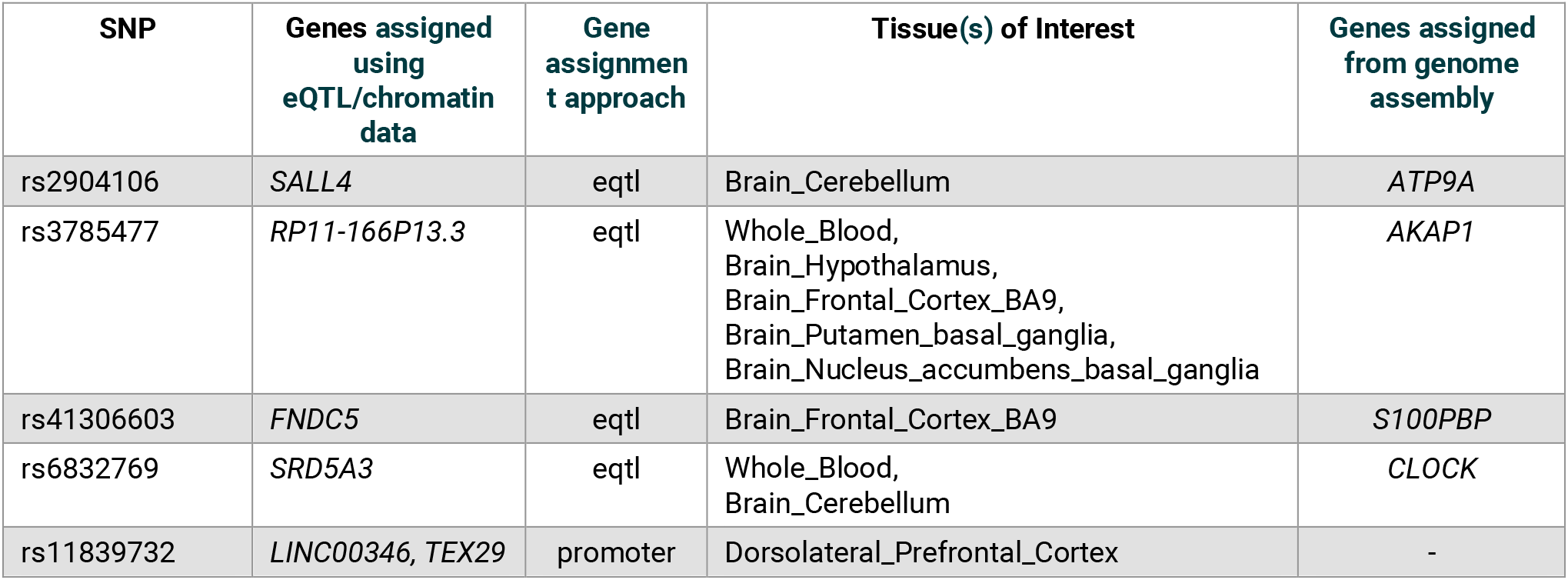
Gene assignments for SNPs using publicly available eQTL and chromatin-interaction data.

**Table 15:** Example public annotation sources for various types of study results

| Annotation Type | Data Source(s) |
| --- | --- |
| Encoded Protein Structure and Function | Uniprot, InterPro, PDBe-KB |
| SNP-Disease Association | GWAS Catalog, PharmGKB, OpenTargets |
| Gene-Disease Association | PubMed, PubChem NCBI, OpenTargets |
| Gene Tissue/Cell Expression | Human Protein Atlas, GTEx, Promoter Capture HiC, Expression Atlas |
| Pathways, Interactions or MoA | Reactome, KEGG, Gene Ontology, PANTHER, WikiPathways, STRING, PubMed, IntAct |
| Safety / Toxicology | International Mouse Phenotyping Consortium, Tox21, eTox, HeCaToS, Open Targets |
| Drugs and chemical compounds | ChEMBL, DrugBank, PubChem NCBI, ProbeMiner, PDBe-KB |

**Table 16:**
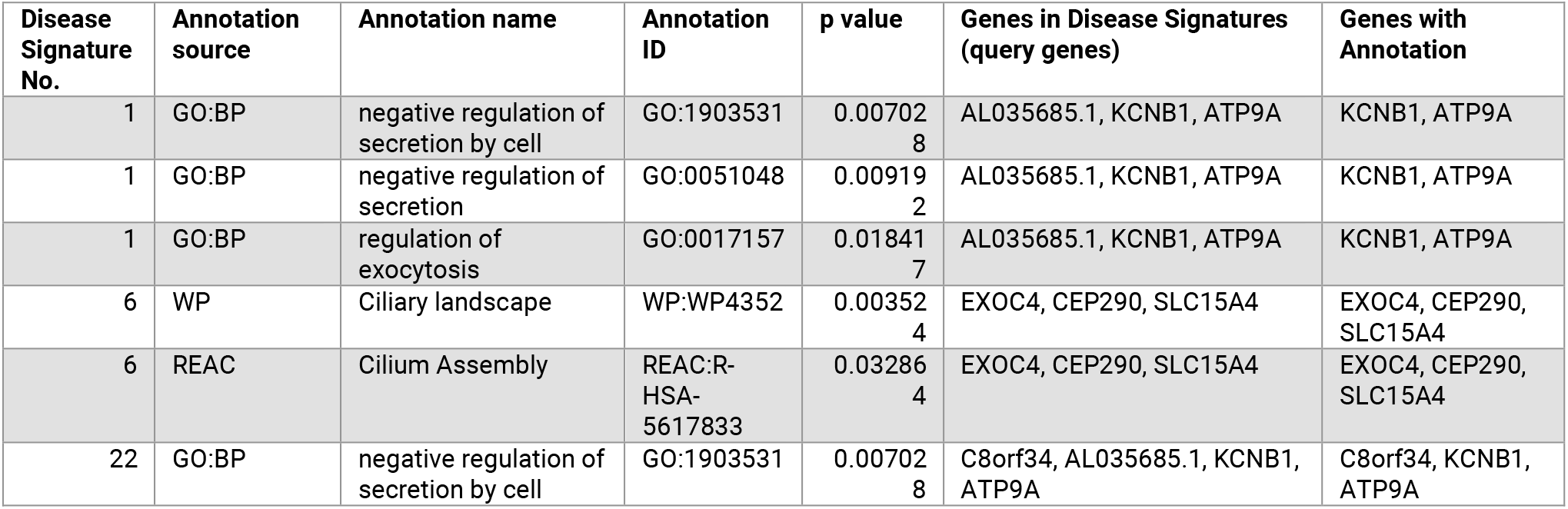
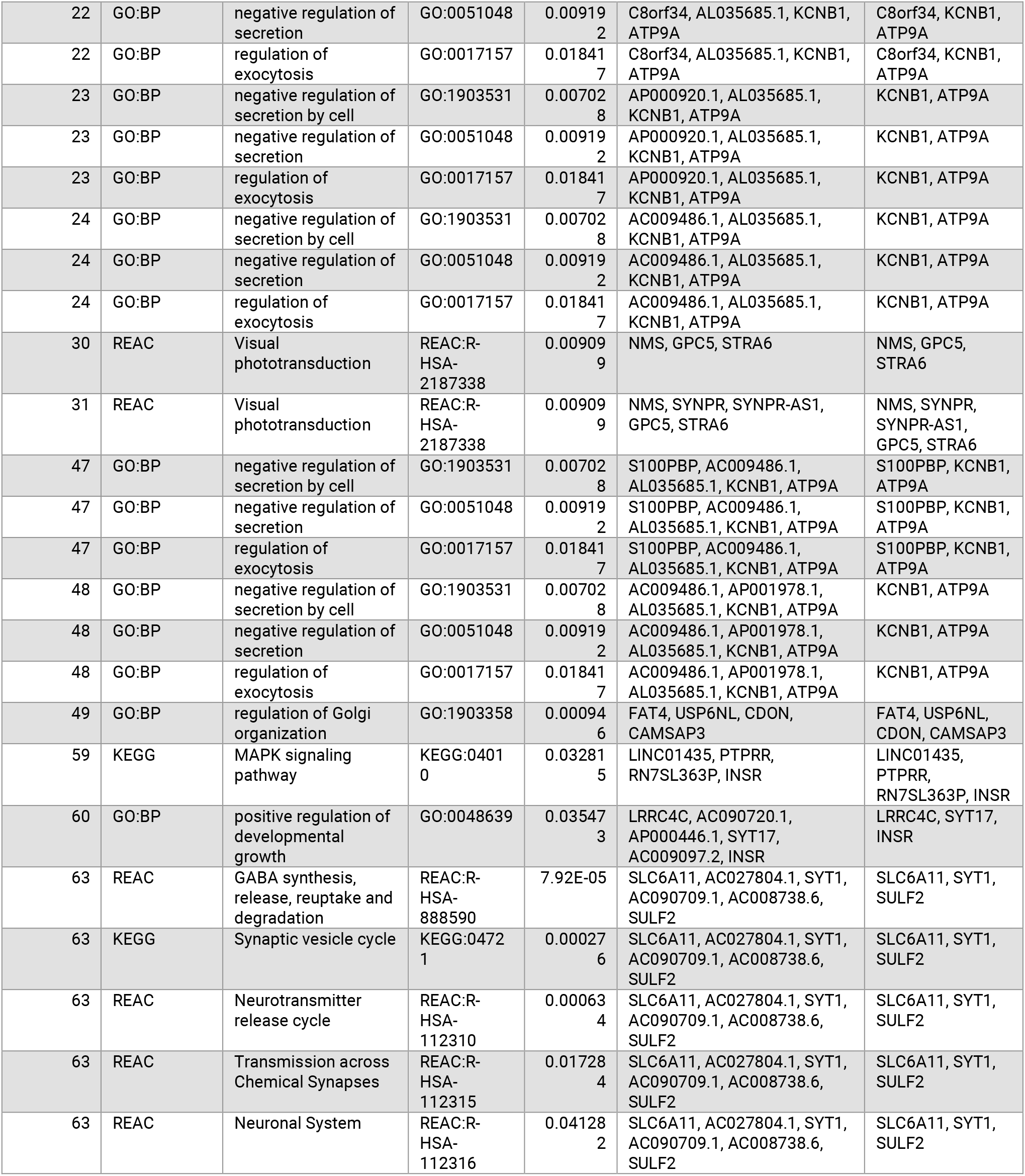
Biological pathway enrichment results for genes associated with disease signatures identified in the ME/CFS Pain Questionnaire study using g:Profiler. Only significant results are reported from the enrichment analysis that used only non-electronic gene annotations, Bonferroni multiple testing correction and considered genes with at least one annotation as background genes.

| Disease Signature No. | Annotation source | Annotation name | Annotation ID | p value | Genes in Disease Signatures (query genes) | Genes with Annotation |
| --- | --- | --- | --- | --- | --- | --- |
| 1 | GO:BP | negative regulation of secretion by cell | GO:1903531 | 0.007028 | AL035685.1, KCNB1, ATP9A | KCNB1, ATP9A |
| 1 | GO:BP | negative regulation of secretion | GO:0051048 | 0.009192 | AL035685.1, KCNB1, ATP9A | KCNB1, ATP9A |
| 1 | GO:BP | regulation of exocytosis | GO:0017157 | 0.018417 | AL035685.1, KCNB1, ATP9A | KCNB1, ATP9A |
| 6 | WP | Ciliary landscape | WP:WP4352 | 0.003524 | EXOC4, CEP290, SLC15A4 | EXOC4, CEP290, SLC15A4 |
| 6 | REAC | Cilium Assembly | REAC:R-HSA-5617833 | 0.032864 | EXOC4, CEP290, SLC15A4 | EXOC4, CEP290, SLC15A4 |
| 22 | GO:BP | negative regulation of secretion by cell | GO:1903531 | 0.007028 | C8orf34, AL035685.1, KCNB1, ATP9A | C8orf34, KCNB1, ATP9A |
| 22 | GO:BP | negative regulation of secretion | GO:0051048 | 0.00919<br>2 | C8orf34, AL035685.1, KCNB1, ATP9A | C8orf34, KCNB1, ATP9A |
| 22 | GO:BP | regulation of exocytosis | GO:0017157 | 0.01841<br>7 | C8orf34, AL035685.1, KCNB1, ATP9A | C8orf34, KCNB1, ATP9A |
| 23 | GO:BP | negative regulation of secretion by cell | GO:1903531 | 0.00702<br>8 | AP000920.1, AL035685.1, KCNB1, ATP9A | KCNB1, ATP9A |
| 23 | GO:BP | negative regulation of secretion | GO:0051048 | 0.00919<br>2 | AP000920.1, AL035685.1, KCNB1, ATP9A | KCNB1, ATP9A |
| 23 | GO:BP | regulation of exocytosis | GO:0017157 | 0.01841<br>7 | AP000920.1, AL035685.1, KCNB1, ATP9A | KCNB1, ATP9A |
| 24 | GO:BP | negative regulation of secretion by cell | GO:1903531 | 0.00702<br>8 | AC009486.1, AL035685.1, KCNB1, ATP9A | KCNB1, ATP9A |
| 24 | GO:BP | negative regulation of secretion | GO:0051048 | 0.00919<br>2 | AC009486.1, AL035685.1, KCNB1, ATP9A | KCNB1, ATP9A |
| 24 | GO:BP | regulation of exocytosis | GO:0017157 | 0.01841<br>7 | AC009486.1, AL035685.1, KCNB1, ATP9A | KCNB1, ATP9A |
| 30 | REAC | Visual phototransduction | REAC:R-HSA-2187338 | 0.00909<br>9 | NMS, GPC5, STRA6 | NMS, GPC5, STRA6 |
| 31 | REAC | Visual phototransduction | REAC:R-HSA-2187338 | 0.00909<br>9 | NMS, SYNPR, SYNPR-AS1, GPC5, STRA6 | NMS, SYNPR, SYNPR-AS1, GPC5, STRA6 |
| 47 | GO:BP | negative regulation of secretion by cell | GO:1903531 | 0.00702<br>8 | S100PBP, AC009486.1, AL035685.1, KCNB1, ATP9A | S100PBP, KCNB1, ATP9A |
| 47 | GO:BP | negative regulation of secretion | GO:0051048 | 0.00919<br>2 | S100PBP, AC009486.1, AL035685.1, KCNB1, ATP9A | S100PBP, KCNB1, ATP9A |
| 47 | GO:BP | regulation of exocytosis | GO:0017157 | 0.01841<br>7 | S100PBP, AC009486.1, AL035685.1, KCNB1, ATP9A | S100PBP, KCNB1, ATP9A |
| 48 | GO:BP | negative regulation of secretion by cell | GO:1903531 | 0.00702<br>8 | AC009486.1, AP001978.1, AL035685.1, KCNB1, ATP9A | KCNB1, ATP9A |
| 48 | GO:BP | negative regulation of secretion | GO:0051048 | 0.00919<br>2 | AC009486.1, AP001978.1, AL035685.1, KCNB1, ATP9A | KCNB1, ATP9A |
| 48 | GO:BP | regulation of exocytosis | GO:0017157 | 0.01841<br>7 | AC009486.1, AP001978.1, AL035685.1, KCNB1, ATP9A | KCNB1, ATP9A |
| 49 | GO:BP | regulation of Golgi organization | GO:1903358 | 0.00094<br>6 | FAT4, USP6NL, CDON, CAMSAP3 | FAT4, USP6NL, CDON, CAMSAP3 |
| 59 | KEGG | MAPK signaling pathway | KEGG:04010 | 0.03281<br>5 | LINC01435, PTPRR, RN7SL363P, INSR | LINC01435, PTPRR, RN7SL363P, INSR |
| 60 | GO:BP | positive regulation of developmental growth | GO:0048639 | 0.03547<br>3 | LRRC4C, AC090720.1, AP000446.1, SYT17, AC009097.2, INSR | LRRC4C, SYT17, INSR |
| 63 | REAC | GABA synthesis, release, reuptake and degradation | REAC:R-HSA-888590 | 7.92E-05 | SLC6A11, AC027804.1, SYT1, AC090709.1, AC008738.6, SULF2 | SLC6A11, SYT1, SULF2 |
| 63 | KEGG | Synaptic vesicle cycle | KEGG:04721 | 0.00027<br>6 | SLC6A11, AC027804.1, SYT1, AC090709.1, AC008738.6, SULF2 | SLC6A11, SYT1, SULF2 |
| 63 | REAC | Neurotransmitter release cycle | REAC:R-HSA-112310 | 0.00063<br>4 | SLC6A11, AC027804.1, SYT1, AC090709.1, AC008738.6, SULF2 | SLC6A11, SYT1, SULF2 |
| 63 | REAC | Transmission across Chemical Synapses | REAC:R-HSA-112315 | 0.01728<br>4 | SLC6A11, AC027804.1, SYT1, AC090709.1, AC008738.6, SULF2 | SLC6A11, SYT1, SULF2 |
| 63 | REAC | Neuronal System | REAC:R-HSA-112316 | 0.04128<br>2 | SLC6A11, AC027804.1, SYT1, AC090709.1, AC008738.6, SULF2 | SLC6A11, SYT1, SULF2 |

### Replication in Three Disjoint Fatigue-associated Cohorts

#### Disjoint CFS (Verbal Interview) Cohort

We analyzed the disjoint UK Biobank CFS Verbal Interview cohort (1,273 cases, excluding individuals that were common to the Pain Questionnaire cohort, and 4,137 controls) using GWAS and combinatorial analysis.

No SNPs were reported to be significant (*p*<5 × 10^−8^) for the Verbal Interview cohort in a standard GWAS case-control association analysis (Figure 8), however, the genomic loci showing modest association values around *p*=1 × 10^−5^ were found to be different than for the Pain Questionnaire cohort (Figure 3). This is likely due to the low power of the two GWAS, and could mean either that these are not true associations or, less likely, that the two populations simply yield different sets of true associations.

Comparison of the results validating the 199 SNPs from the Pain Questionnaire cohort in the Verbal Interview cohort showed that five (rs2304725, rs2904106, rs9444564, rs10420798 and rs11695478) of the 25 critical SNPs identified in the Pain Questionnaire cohort were replicated in this analysis. Two of these critical SNPs mapped to the *SLC6A11* (rs2304725) and *ATP9A* (rs2904106) genes, which were identified in this cohort. This suggests that these five critical SNPs (and two genes) are particularly strongly associated with ME/CFS. None of the replicated SNPs has significant direct GWAS associations to the disease or traits.

#### Disjoint Post-Viral Syndrome Cohort

Three critical SNPs (rs56218501/Affx-16805420, rs12530627 and rs2499908) identified in the Pain Questionnaire cohort were replicated in the fibromyalgia cohort analysis. Two of these critical SNPs mapped to the genes SULF2 (rs56218501/Affx-16805420) and USP6NL (rs2499908).

#### Disjoint Fibromyalgia Cohort

Two critical SNPs (rs10420798 and rs2499908) and the gene USP6NL (mapped to rs2499908) identified in the Pain Questionnaire cohort were replicated in the fibromyalgia cohort analysis. This suggests that these two critical SNPs and the gene USP6NL are particularly strongly associated with fibromyalgia cases.

### Disease Mechanisms and Genetic Functions

We used a detailed analysis of the metabolic context, exploiting an integrated semantic knowledge graph drawing from different data sources including Open Targets^110^ associations, known gene-disease associations from scientific literature, mouse phenotypes etc., to annotate the 14 genes identified in the analysis of the Pain Questionnaire cohort.

While acknowledging annotation bias and an inevitable degree of subjectivity, we applied consistent heuristics to the available knowledge around a target, enabling us to identify that variants in these genes might impact different cellular processes. The five cellular processes or biological systems identified have previously been associated with ME/CFS – namely, susceptibility to infection, autoimmune and chronic inflammation development, metabolic dysfunction, increased vulnerability to stress and sleep disturbance – and it is possible to form plausible disease phenotype hypotheses for them (Table 6).

Furthermore, critical SNPs found in the same patient community may be mapped to genes with shared biological functions or pathways. Pathway enrichment analysis of the genes using gprofiler^57^ (excluding electronic Gene Ontology annotations), indicated that two large communities identified in this analysis containing multiple critical SNPs – Community 1 and Community 15 (Table 5) – may be implicated in common biological processes (Supplementary Table 14).

Community 1 contains three critical SNPs; rs41306603, a 3 prime UTR variant mapping to *S100PBP*, and two intronic variants, rs2904106 and rs237475, found in *ATP9A* and *KCNB1* respectively. The genetic variants in *ATP9A* and *KCNB1* were found in the same disease signature (an example is shown in Table 3), which shows significant enrichment linked to regulation of exocytosis and negative regulation of secretion by cell (GO annotations, Supplementary Table 16). Using additional evidence from the scientific literature, both *ATP9A* and *KCNB1* are expressed in pancreatic beta cells and are involved in the regulation of insulin secretion (Table 6)^36, 38^. This could suggest a combined biological effect of these two co-associated SNPs in causing dysregulated insulin signalling in this subgroup of ME/CFS patients.

Community 15 contains two critical SNPs; rs2304725, a synonymous variant in *SLC6A11*, and rs56218501/ Affx-16805420, a missense variant in *SULF2*. This disease signature shows enrichment for GABA synthesis and release and synaptic vesicle cycle (Reactome and KEGG annotations, Supplementary Table 16). This enrichment can be supported by further literature evidence that indicates the association of these genes with CNS-related pathways (Table 6)^56, 58, 59^.

Many of the patient communities identified contain genes that could be categorized into more than one of these mechanisms and there was no clear distinction in biological pathways when communities were compared (Figure 9). This supports the hypothesis that development of ME/CFS is caused by the interaction and subsequent dysregulation of multiple immune, metabolic and neuronal pathways in combination.

We used the additional phenotypic and clinical data available in the UK Biobank to generate a patient profile for each patient community. However, the validation and significance of these findings are limited by the scope and depth of disease related data collected in UK Biobank (and other sources) that is available and relevant to ME/CFS patients, and the paucity of disease models.

#### Viral/Bacterial Susceptibility

ME/CFS onset is often thought be linked to viral infection in patients, although no specific single viral or bacterial trigger has yet been confirmed^58^. There have been reports of shared pathophysiological, clinical, and transcriptomic features between viral and/or bacterial diseases and ME/CFS^59,60^.

We identified five genes – *S100PBP, AKAP1, USP6NL, CDON* and *SULF2* – in five different patient subgroups that have been associated with viral and/or bacterial infection in the literature (Table 6).

These may represent a subset of ME/CFS patients with increased susceptibility to infection, or differential response to infection that leads to ineffectual viral clearance. We therefore evaluated the clinical records of the ME/CFS case population included in this study to identify any evidence of prior infection of the most common ME/CFS-associated infective triggers, including infectious mononucleosis, and EBV and/or Herpesviruses’ seropositivity. Unfortunately, the total numbers of patients and clinical reports with any of these was too small (approximately 2% of cases) to generate any statistically significant gene/patient subgroup associations, so the question of whether any such significant associations exist remains unanswered.

#### Autoimmune and Chronic Inflammation

Our analysis identified seven genes that have been associated with diseases that have autoimmune components in both the literature and in other disease studies that we have undertaken, including COVID-19, rheumatoid arthritis and Sjögren’s syndrome (unpublished results).

ME/CFS shares several characteristics with autoimmune diseases, including the increased level of pro-inflammatory cytokines and higher prevalence in females, with as many as 60% of ME/CFS patients also reported to be diagnosed with an autoimmune disease^61,62^.This co-association with other autoimmune diseases was also evident in our analysis of ME/CFS patients when compared against the rest of the UK Biobank population (Figure 2). Whether this reflects a real association or misdiagnosis of patients remains unclear.

We speculate that increased susceptibility to viral infection in ME/CFS patients, resulting in recurrent or chronic infections, may also drive chronic inflammation and autoimmune development.^63^ Furthermore, pro-inflammatory cytokines associated with autoimmune development have also been shown to contribute to mitochondrial dysfunction and decreased respiratory capacity, and there is evidence that patients with other autoimmune diseases also display mitochondrial dysfunction^64,65,66^.

Solute carrier family member 15 (SLC15A4) is found in the lysosomal membrane and has enriched expression in immune cells. Genetic variants in *SLC15A4* have been associated with increased risk of developing inflammatory diseases like systemic lupus erythematosus.^67^ Interestingly, SLC15A4 has been shown to play a crucial role in immune cell tolerance to metabolic stress via AMPK and mTORC1 and maintenance of respiratory homeostasis in innate immune cells^68^ and *SLC15A4* knock down results in decreased mitochondrial function under cell stress^69^. No eQTL associations were found for the ME/CFS SNP linked to *SLC15A4*.

A specific variant – associated to *GPC5* (glypican 5) – was found in 17% (408) of ME/CFS cases in the Pain Questionnaire study. Glypican 5 is a cell surface proteoglycan that has been identified in many different multiple sclerosis genetic studies^70,71,72^. A further four – *ATP9A, TMEM232, PHACTR2* and *SLC6A11 -* out of the seven autoimmune genes identified in this study can also be linked to multiple sclerosis development (Table 6). MS and ME/CFS are believed to have a viral trigger component, such as Epstein-Barr virus (EBV), and their patients share similar symptoms, including fatigue, pain, sleep disturbance and cognitive dysfunction^73^.

#### Metabolic Dysfunction

Reductions in reserve capacity and inability to raise mitochondrial respiration in response to stress compared with controls indicates that ME/CFS patients are less able to meet energy demands, resulting in increased fatigue and exercise intolerance^74^.

Combinations of genes including a variant in *AKAP1* were found in 27% (648) of the Pain Questionnaire cases – the highest proportion for any RF-scored genetic variant identified in the study – and no more than 1.5% of controls. AKAP1 (A kinase (PRKA) anchor protein 1) is a scaffold protein in the mitochondrial membrane, regulating mitochondrial respiration via AMPK. A study has demonstrated that phosphorylation of AKAP1 by AMPK was crucial for AMPK-induced increase in mitochondrial respiration in human muscle post-exercise.^75^ Furthermore, knockout of *AKAP1* in mice resulted in reduced skeletal muscle capillary density and functional recovery impairment in addition to increased mitochondrial dysfunction and cellular stress in endothelial cells^76^. The identification of a disease associated *AKAP1* variant in this study provides a strong genetic link to mitochondrial dysfunction and the reduction in energy capacity observed in biochemical analysis of ME/CFS patients^77^.

We also identified a series of genes involved in other metabolic processes such as insulin sensitivity and lipid metabolism.

ATP9A is a member of the Type IV P-type ATPases (P4-ATPases) family involved in the process of lipid flipping. ATP9A may regulate intracellular levels of ceramide and sphingosine^78^, which have been shown to be altered in patients with chronic fatigue and in the skeletal muscle of fatigue-associated conditions^79,80,81^.

ATP9A is also expressed in pancreatic beta cells and has a role in driving glucose-stimulated insulin release^82^. Moreover, a variant in *ATP9A* has been associated with multiple sclerosis in a homozygosity haplotype analysis^83^.

Finally, 348 (15%) ME/CFS patients (and 4% of controls) from this study were most associated with the community of genetic variants including the gene encoding the insulin receptor (*INSR*). A study has found that insulin levels in ME/CFS patients were higher than in healthy controls^84^, which is hypothesized to be as a results of insulin resistance and ischemia-reperfusion damage in skeletal muscles of patients with ME/CFS^85^.

These 348 ME/CFS patients also presented with relatively higher blood levels of lactate from the UK Biobank NMR metabolomics data (*p*<0.017, Supplementary Table 13) compared to the entire case population. Lactate is implicated in insulin resistance, resulting in reduced insulin-dependent glucose uptake in skeletal muscle and dysregulated insulin signaling^86,87^. Dysregulation of insulin and lactate in ME/CFS patients may also have an impact on mitochondrial function, decreasing mitochondrial size and respiratory function^88^.

#### Response to Stress

Three of the genetic variants that were significant in the Pain Questionnaire analysis – located in genes *SLC6A11, SULF2* and *CDON* – were identified in communities of ME/CFS patients more likely (*p*=0.003, Supplementary Table 12) to report the occurrence of illness and psychosocial factors (injury, bereavement, stress) in the last 3-8 years. These could represent a subset of ME/CFS patients with combinations of variants involving these genes that confer vulnerability to psychological stress.

SLC6A11 (GAT3) is a sodium-dependent transporter involved in GABA reuptake at presynaptic terminals. Altered levels of GAT3 have been associated with increased neuroinflammation and cognitive impairment^89^, in addition to sleep disturbance, juvenile stress and depression in animal models^90,91,92,93^. Furthermore, patients with SNP combinations including those in *SLC6A11* show increased levels of phenylalanine (*p*=0.022) in the metabolomics data compared to other ME/CFS subgroups identified in our analysis (although this is not significant after multiple testing correction). Phenylalanine is a precursor for monoamine neurotransmitters, such as dopamine, epinephrine and serotonin. Finally, two further SNPs in *SLC6A11* were also identified to be significant in the Verbal Interview ME/CFS case dataset, providing additional evidence for the importance of this gene in ME/CFS development.

Sulfatase 2 (SULF2) is an enzyme that regulates the effects of heparan sulfate. A variant in this gene is found in combinations with *SLC6A11* and is therefore also associated with the patient community with raised phenylalanine levels. Sulfatase 2 plays a role in a wide variety of biological process and is expressed in most tissues (Figure 16 in Supplementary Data). Sulfatase 2 is crucial for brain development, contributing to processes such as neurite outgrowth and responsiveness to growth factors^94,95,96^, and there is an association between *SULF2* variants and HSV-1 and depression risk, and also with malaise and fatigue in UK Biobank studies^97,98^. The directionality of the specific observed SULF2 association, i.e. whether this is a ‘risk’ allele or a ‘protective’ allele, is not however clear from this study or the literature. Although there is a known association with sex hormone globulin levels, we did not find any difference in male:female distribution for this community.

*CDON* (cell adhesion associated, oncogene regulated) encodes a cell surface receptor that is highly involved in muscle regeneration^99^. In muscle cells, depletion of CDON results in impaired muscle regeneration and senescence, as well as increased cell stress^100^. However, CDON has also been associated with complicated bacteremia^101^ and development of midbrain dopamine pathways^102^. This indicates that CDON has a diverse range of functional roles that could impact ME/CFS development.

#### Sleep Disturbance

We identified two genes that could play a role in the sleep disturbance often reported by ME/CFS patients, *SLC6A11* and *CLOCK*. The *CLOCK* (Circadian Locomotor Output Cycles Kaput) gene is one of the key regulators of circadian rhythm. Altered circadian rhythm is hypothesized to contribute to many of the symptoms experienced by patients with ME/CFS, including insomnia, pain and post-exertional malaise.^103^ This is because disruptions in the circadian clock have far reaching biological consequences beyond sleep disruption, including disturbed mitochondrial function, dysregulated cellular stress responses and insulin sensitivity^104,105,106^. Furthermore, transcriptomic analysis of peripheral blood mononuclear cells indicated that several genes involved in circadian rhythm were elevated in ME/CFS patients^107^.

We also found significant enrichment in patients with variants in *CLOCK* who also had been diagnosed with fibromyalgia. ME/CFS and fibromyalgia patients exhibit similar symptoms, including fatigue, cognitive functioning impairment and pain, which could indicate similar underlying biological drivers of disease (or a degree of misdiagnosis). Interestingly, a study investigating the differences between the two conditions found that patients with both ME/CFS and fibromyalgia also presented with sleep disruption, in contrast to CFS only patients and healthy controls^108^. These results could reveal further insights into the cause of this symptom.

### Drug Target Evaluation

There are currently no specific pharmacological treatment options for ME/CFS patients. The detailed insights generated by combinatorial analysis of this UK Biobank population can be used to inform the development of novel drug targets guided by patient stratification biomarkers associated with each of the ME/CFS subgroups.

In other studies, for example in motor neuron disease / amyotrophic lateral sclerosis (ALS), we have at this stage identified known pharmacological modulators of several novel targets discovered using the approach described above. We tested these in a patient-derived human induced neuronal progenitor cells (iNPC) cellular assay with a co-culture of motor neurons, microglia and astrocytes^109^ to provide biological validation of the disease modification potential of modulating several novel targets identified using this methodology (manuscript in preparation). We are further developing direct CRISPR derived knock-in/knock-outs for those targets in iPSC-derived neurons. However, in ME/CFS, not only are there no assays or model systems available, but we also do not understand the tissues involved in various aspects of the disease. This prevents the ready evaluation of the effects of modulating the targets either pharmacologically or via direct genetic manipulation.

Each gene identified in this study was nonetheless evaluated for drug tractability (Table 8), indicating that seven of the targets exhibit potential small molecule or antibody tractability. Moreover, three of the genes are targeted by drugs in clinical development, suggesting their potential as drug repositioning candidates, which might offer a faster and derisked route to approval if their safety and efficacy can be demonstrated.

## Discussion

After decades of study, the genetic contributions to the etiology of ME/CFS and the different mechanisms underpinning the disease remain poorly understood. It is unsurprising therefore that our analysis demonstrates that ME/CFS at a genetic level is polygenic and heterogeneous.

This is confirmed both by the genetic association and patient stratification results generated using combinatorial analysis techniques in this study, as well as the consistent failure of previous GWAS analyses to find replicable signal within this cohort and/or between ME/CFS population datasets, which would be expected if clinically relevant monogenic signals were present^6^.

Using a hypothesis-free combinatorial analytics approach based on the PrecisionLife platform, we identified 199 SNPs in 84 high-order combinations that were highly associated with 91% of the ME/CFS cases in the UK Biobank Pain Questionnaire cohort. These variants could be mapped to 14 genes, which appear to be compatible with the major cellular mechanisms suspected by other groups working in the field (ref) and show a level of overlap with diseases sharing similar symptoms, such as MS^111^and long Covid^112,113^.

We further used these findings to stratify the ME/CFS patients genetically and correlated this stratification with clinical criteria. There is a degree of evidence of replication of several SNPs and two of those genes being identified in a second UK Biobank cohort, and the consistency of results from internal cross-validation replication runs is also encouraging.

Biological analysis of these genes indicates that many of them are directly linked to the key cellular mechanisms hypothesized to underpin ME/CFS, including vulnerabilities to stress and infection, mitochondrial dysfunction, sleep disturbance and autoimmune development. This has revealed several potential novel drug targets that could be the basis of targeted therapy development for ME/CFS patients.

### Study Limitations

There are however a number of limitations with this study. Analysis of ME/CFS data is complicated by several logistical factors impacting data availability and quality, including low reporting rates, inaccurate diagnosis, limited cohorts with genetic information, and limited longitudinal clinical, psychosocial, epidemiological, and environmental data. This is exacerbated by the nature of the disease with its complex interactions of multiple etiologies, mechanisms, and influences.

The UK Biobank cohort, while essential to enabling this analysis, represents only a small cohort of atypically older ME/CFS patients with predominantly white, European ancestry who have self-reported their clinical diagnosis. The lack of detailed ME/CFS-specific supporting clinical and/or phenotypic data makes it hard to evaluate individual clinical experiences and assess potential triggers of disease onset, recovery or relapse.

While we have tried to replicate the analysis and results between two different ME/CFS UK Biobank cohorts, a high rate of false negatives, the self-reporting of the clinical diagnosis, which in some cases may be misdiagnosed, and other variations in the case criteria between the cohorts make expectation of a complete correlation of results unrealistic. It is nonetheless encouraging that five critical SNPs and two of the genes identified do in fact appear in both cohorts, even allowing for the shared genetic ancestries of the cohorts.

Although it can occur at any time of life, the average age at onset of ME/CFS is in the 30s^114^, perhaps with an earlier secondary peak^115^, whereas the average age of the UK Biobank population is 56 years^116^ and the population has a selective participation bias to ‘healthy volunteers’^117^. In the Pain Questionnaire study, the average age of cases was 69 years, indicating an even greater bias to a more elderly population. This might cause the associations identified to be skewed away from causes that could be more prevalent in a more age inclusive population or towards comorbidities that exerted a larger influence. On the other hand, an older population may be more accurately diagnosed. A better distribution of ages and longitudinal follow-up data would enable analysis of differences in etiology, clinical presentation or comorbidities and prescriptions.

ME/CFS is clearly a complex disease with multiple endogenous and exogenous triggers, potentially ranging from metabolism, autoimmune and infection, to stress and environmental impacts. Not all of these factors are recorded consistently and accurately in the available dataset, making their influence across one of more of the patient subgroups hard to determine definitively.

Finally, there is a considerable bias in the makeup of the patients both in UK Biobank and in this study. All of the participants in this study have a European ancestry due to their predominance in the source data^22^. There may well be different and additional mechanisms influencing the disease in cohorts with other ancestries and geographies (including different triggering pathogens).

### Similarities with other Diseases

MS and ME/CFS patients share a number of similar symptoms, including pain, sleep disturbance and cognitive dysfunction^118^, and both can have a viral trigger such as Epstein-Barr virus (EBV)^4,119^. There is also increasing evidence that many patients diagnosed with long COVID share similar symptoms, such as chronic fatigue and ‘brain fog’, with individuals with ME/CFS. It is also believed that some patients may be developing ME/CFS as a direct result of having a COVID-19 infection^120,121,122^.

This suggests that the two diseases may share similar etiologies with possible overlap in the biological drivers and risk genes. Our analysis of the first UK Biobank COVID-19 population identified four genes out of 68 associated specifically with the risk of severe COVID that we had previously identified as having strong association with neurodegenerative processes^23^, including *ATXN1, SORCS2* and *STH* and *MAPT* from loci on chromosome 17 that were subsequently validated by the results from the COVID-19 Host Genetics Initiative^123^. This analysis also revealed several other disease and symptom associated mechanisms, such as viral host response factors and pro-inflammatory cytokine production.

We are in the process of analyzing two populations in long COVID-19 (Sano Genetics, GOLD study) and multiple sclerosis (UK Biobank) in order to identify any shared genes and biological mechanisms underpinning ME/CFS, multiple sclerosis and long COVID-19 development. Preliminary findings from our long COVID analysis have indicated that three of the genes identified in this study are also significant in the long COVID patient group (albeit with different SNPs, but again none of these are in LD). These will be subject of further validation in a new publication later this year.

## Conclusion/Future Perspectives

The use of a hypothesis-free combinatorial analytics approach using the PrecisionLife platform has enabled us to identify 14 novel genetic associations with ME/CFS in a UK Biobank cohort. Several previous attempts at GWAS approaches^12^ have failed to validate single SNP associations or highlight significant risk genes in this ME/CFS cohort.

This study has produced further evidence of the polygenic and heterogeneous nature of the disease and produced patient stratification results that describe the mechanistic etiology of the disease. This also suggests a set of novel potential drug targets that may be relevant for the major ME/CFS patient subgroups.

There are a number of limitations with this study discussed above, and a larger, more detailed longitudinal patient dataset is likely to significantly improve the results. For this reason, we aim to replicate and extend the results from this UK Biobank study with combinatorial analysis of a future DecodeME study. DecodeME is the largest current genetic ME/CFS study, with over 20,000 participants involved^124^, and the more detailed patient survey data collected is likely to allow deeper insights into the different subgroups and targets involved with the disease.

The findings of this study nonetheless provide some indicators of useful areas of study in terms of diagnostics, novel drug targets, and potentially precision repositioning opportunities. As a first step, simply identifying and validating patient stratification biomarkers that could be used to create an accurate risk model or diagnostic test for ME/CFS would be a huge step forward in recognition and treatment of the disease.

Discovery of drug candidates for ME/CFS has been limited in progress not just due to lack of plausible targets (and disease involved tissues), but also access to accurate models of the various aspects of the disease. Biological validation of the disease modification potential of the identified targets *in vitro* or *in vivo* is the next obvious step, but the lack of ready access to validated assays and disease models, or even a specific cell type to target is a barrier.

We hope that with a smaller set of genes on which to focus, genetic interventions (e.g., CRISPR knock in/out) or transient siRNA modulation might enable us to generate cell lines that capture features of the disease biology and to investigate in a cellular system the role that each target gene plays. We could further use these modified/modulated cell lines as assays to evaluate recovery of a normal phenotype in the presence of active molecules to accelerate the discovery and validation of novel and/or precision repositioned therapeutics.

We have identified known active compounds acting at three of the targets found in this study using precision repositioning approaches^125^, and there is the potential to evaluate the likely impact of these retrospectively via analysis of real-world data collections with longitudinal prescription information, and also pharmacologically in the new assay systems using known active drugs and/or development candidates as tool compounds. Given a good safety profile for these compounds or their derivatives, this may provide sufficient evidence in the future for the design of first in man studies.

Finally, understanding the drivers of ME/CFS and disorders with similar symptoms such as long COVID and MS, and establishing the similarities and differences between them in more detail is likely to have profound implications for patients. Accurate diagnosis and effective treatment options are limited in all of these diseases, and we hope that uncovering of the disease etiologies, better patient stratification, and identification of novel drug targets will yield rapid progress in approval of better diagnostic tools and drugs for patients.

## Data Availability

All data sources are described in the Supplementary Information, and no new source data were collected. Only data from existing UK Biobank study cohorts were analyzed. All datasets generated during the study are described in the Supplementary Data section and/or available from the corresponding author upon reasonable request.

## Declarations

### Ethics Approval

Research described in this article has been conducted using data from UK Biobank Resource (application number 44288). UK Biobank has approval from the North West Multi-centre Research Ethics Committee (MREC) as a Research Tissue Bank (RTB) approval, and researchers do not require separate ethical clearance and can operate under this RTB approval.

### Consent for Publication

Not applicable

### Authors’ contributions

S.G., S.D., and K.T. wrote the manuscript. S.G. designed the approach, and K.T., S.D., J.K., and J.S. analyzed the data generated by the PrecisionLife platform. All authors provided input and approved the final version of the manuscript.

## Acknowledgements

Research described in this article has been conducted using data from UK Biobank Resource (application number 44288). We would like to particularly acknowledge the helpful advice and encouragement provided by Prof. Chris Ponting, MRC Investigator at the MRC Human Genetics Unit, Institute of Genetics and Cancer of the University of Edinburgh and Sonya Chowdhury, CEO of Action for ME. Special thanks to Matthew Pearson, Karan Dahele, and Mark Strivens who provided input into the manuscript, Gert Møller, who initially developed the combinatorial analytics methodology, and the rest of the PrecisionLife team.

## Supplementary Data

### Pain Questionnaire Study Design

UK Biobank participants included in the Pain Questionnaire case cohort answered positively to the question:

| Data Type | ICD-10 / Data Field ID | Description |
| --- | --- | --- |
| Self-reported | 120010 | Ever had Chronic Fatigue Syndrome or Myalgic Encephalitis (M.E.) |

UK Biobank participants excluded from the Pain Questionnaire control cohort met one or more of the following criteria:

### Genotype Quality Control

Appropriate quality control of genotype data was performed using PLINK^29^ and GRAF^126^ (Genetic Relationship and Fingerprinting) based on standard quality control procedures to ensure thorough cleaning of the data before it is used for genomic analyses.

This included the following steps:

1. Batch effect correction: Batch-level quality control procedures was performed based on recommendations by UK Biobank^22^ and only SNPs that pass all batch-level QC were used for further analysis.
2. Sample and SNP filtering based on missingness: The filtering for SNPs with missing data (<5%) was followed by filtering of individuals with missing data (<5%) using PLINK.
3. Minor Allele Frequency (MAF) filtering of SNPs: The genotype data would be filtered to exclude SNPs with MAF <0.0001 using PLINK.
4. Hardy-Weinberg Equilibrium (HWE) filtering: HWE filtering was performed on controls with *p*<10^−10^ using PLINK.
5. Heterozygosity filtering: Samples with extreme (very high or very low) heterozygosity were removed.
6. Sample filtering based on relatedness: GRAF-rel^127^ was used to identify duplicates and closely related subjects in the dataset. After identification of close relatives, only one representative of each closely related family pairs was retained.
7. Ancestry analysis: GRAF-pop was used for ancestry inference and limit samples for the dataset to European ancestry.
8. Sex discrepancy of individuals: Samples that have discrepancies between the sex recorded in the dataset and their sex based on absence/presence of a Y chromosome were removed.

### Cohort Analysis

#### Data used for Cohort Analysis

Data used for cohort analysis in Figure 2:

##### 1. Exposure to infectious agents

1. Exposure to infectious agents:
| Data Type | ICD-10/ Data Field /Category ID | Description |
| --- | --- | --- |
| Blood assays | 23053 | EBV seropositivity for Epstein-Barr Virus |
| Blood assays | 1307 | Infectious disease antigens |
| HES | A00.x-B99.x | Chapter I Certain infectious and parasitic diseases |

##### 2. Diagnosis with any of the most common autoimmune diseases

2. Diagnosis with any of the most common autoimmune diseases:
| Data Type | ICD-10 / Data Field / Category ID | Description |
| --- | --- | --- |
| HES | M05.x or M06.x | Rheumatoid arthritis |
| HES | L40.x or M07.x | Psoriasis / psoriatic arthritis |
| HES | G35.x | Multiple sclerosis |
| HES | M32.x | Systemic lupus erythematosus |
| HES | K50.x or K51.x | Inflammatory bowel disease (Crohn's and ulcerative colitis) |
| HES | K90.0 | Celiac disease |
| HES | M45.x | Ankylosing spondylitis |
| HES | M35.0 | Sjogren's syndrome |
| HES | M35.3 | Polymyalgia rheumatica |
| HES | M34.x | Systemic sclerosis |
| HES | M79.7 | Fibromyalgia |
| HES | E10.x | Type 1 diabetes |
| HES | E06.x | Thyroiditis |

##### 3. Evidence of significant stressful events

3. Evidence of significant stressful events:
| Data Type | ICD-10 / Data Field /Category ID | Description |
| --- | --- | --- |
| Self-reported | 6145 | Illness, injury, bereavement, stress in last two years (any but 'none of the above') |
| Self-reported | 20499 | Ever sought or received professional help for mental distress |
| Self-reported | 20500 | Ever suffered mental distress preventing usual activities |
| HES | Chapter XX | External causes of morbidity and mortality |

#### Sex

#### Age & BMI

### Combinatorial Disease Signatures

As an internal validation we generated five smaller subsets of the Pain Questionnaire cohort each excluding a different 10% of the cases. These cross-validation runs therefore comprised 90% of the case population compared against the same set of controls as the Pain Questionnaire cohort. We compared the odds ratios and Z-scores of the disease signatures identified in the full cohort to those identified in each of the subset analyses. The odds ratios and Z-scores of the SNP combinations remain largely consistent irrespective of small changes to the case population (Figure 15).

This technical (internal cross-validation) replicate study provided an internal parameter check, but it has insufficient independence between both the case and control sets used in the runs for reliable use as formal replication.

### Genes Associated with Phenotypes

### Genes Associated with SNPs using eQTL and Chromatin Interaction Data

### Tissue Expression of Genes

### Comparison with UK Biobank CFS Verbal Interview Study

### Gene Annotation Data Sources

